# Construction and Validation of a Nomogram Based on the Log Odds of Positive Lymph Based on the Log Odds of Positive Lymph Nodes to Predict the Prognosis of Lung Neuroendocrine Tumors

**DOI:** 10.1101/2021.10.12.21264905

**Authors:** Suyu Wang, Juan Wei, Yibin Guo, Qiumeng Xu, Xin Lv, Yue Yu, Meiyun Liu

## Abstract

**Objectives:** This study aimed to investigate the prognostic value of Log odds of positive lymph nodes (LODDS) for predicting the long-term prognosis of patients with node-positive lung neuroendocrine tumors (LNETs).

**Materials and Methods:** We collected 506 eligible patients with resected N1/N2 classification LNETs from the Surveillance, Epidemiology, and End Results (SEER) database between 2004 and 2015. First, we applied the Cox proportional-hazards regression model to evaluate the relationship between LODDS and study endpoints (cancer-specific survival [CSS] and overall survival [OS]) based on the entire cohort. Second, the study cohort was divided into derivation cohort (n=300) and external validation cohort (n=206) based on different geographic regions. Nomograms were constructed and validated based on these two cohorts to predict the 1-, 3- and 5-year survival of patients with LNETs. The accuracy and clinical practicability of nomograms were tested and compared by Harrell’s concordance index (C-index), integrated discrimination improvement (IDI), net reclassification improvement (NRI), calibration plots, and decision curve analyses.

**Results:** The Cox proportional-hazards model showed the high LODDS group (-0.33≤LODDS≤1.14) had significantly higher mortality compared to those in the low LODDS group (-1.44 ≤LODDS<-0.33) for both CSS and OS. In addition, besides LODDS, age at diagnosis, histotype, type of surgery, radiotherapy, and chemotherapy were shown as independent predictors in Cox regression analyses and included in the nomograms. The values of c-index, NRI, and IDI indicated that the established nomogram performed significantly better than the conventional eighth edition of the TNM staging system alone. The calibration plots for predictions of the 1-, 3-, and 5-year OS were in excellent agreement. Decision curve analyses showed that the nomogram had value in terms of clinical application.

**Conclusions:** We created visualized nomograms for CSS and OS of LNET patients, facilitating clinicians to provide highly individualized risk assessment and therapy.

## 1. Introduction

Lung neuroendocrine tumors (LNETs) originate from pulmonary neuroendocrine cells, accounting for approximately 25% of primary lung neoplasms [1]. Owing to the increased lung cancer screening, the annual incidence of LNET has substantially increased, rising from 0.0003% in 1973 to 0.0014% in 2004 in the United States [2–4]. Currently, the 2015 World Health Organization classification has grouped LNETs into four histologic variants based on their histopathologic features: typical carcinoid (TC), atypical carcinoid (AC), large cell neuroendocrine carcinoma (LCNEC), and small cell lung carcinoma (SCLC) [5]. Due to the rarity and morphological heterogeneity of these tumors, there have been limited clinical data available regarding LNETs, thus making their diagnosis, staging, risk assessment, and treatment challenging [1]. Although specific to non-small cell lung cancer (NSCLC), the international American Joint Committee on Cancer/Union for International Cancer Control (AJCC/UICC) tumor-node-metastasis (TNM) staging system has been applied to LNETs [6, 7]. However, several studies have shown an overlapping survival of patients with LNETs, particularly in stages II and III [6–10]. Therefore, further investigation is warranted to optimize the staging system for LNETs.

Lymph node (LN) involvement is a significant prognostic factor for staging and risk stratification. A combination of the number of positive lymph nodes (NPLN), the number of negative lymph nodes, and the anatomic location of LN metastasis have been applied in the staging of many malignancies [11–15]. However, the latest version of AJCC/UICC TNM classification of lung cancer did not take account of any number- or ratio-based LN staging system, which could affect the precision of prognosis evaluation [16, 17]. Log odds of positive lymph nodes (LODDS), defined as the logarithm of the ratio between NPLN and NNLN, is currently being used as a novel prognostic indicator with a strong ability to identify patients with a homogeneous prognosis in many malignant tumors, including NSCLC [18–21]. However, no specific study has focused on its prognostic significance in LNETs till now.

The present study aimed to determine whether LODDS could be utilized to predict the long-term prognosis of patients with node-positive (N1/N2 classification) LNET using the Surveillance, Epidemiology, and End Results (SEER) database. To facilitate clinical use, we also constructed a visualized and online nomogram incorporating LODDS for LNETs.

## 2. Materials and methods

### 2.1 Study design, data source, and ethical statement

We conducted a multi-center retrospective cohort study according to the parts of the methods described in our previous studies [20–22]. We use the Transparent Reporting of a multivariate prediction model for Individual Prediction or Diagnosis (TRIPOD) for reporting [23]. The data of this study were extracted from the SEER 18 registries research database, covering approximately 28% of the population of the United States [24]. Data were extracted using the SEER*Stat version 8.3.5 software. The requirement for informed consent was waived because the study utilized the anonymous data available in the database. In summary, this study complied with the Declaration of Helsinki [25].

### 2.2 Population selection

Data on the patients with lung cancer were obtained from the SEER database. Inclusion criteria were as followed: (1) diagnosed from 2004 to 2015; (2) site recode “ICD-O-3/WHO 2008” restricted to “Lung and Bronchus”; and (3) pathologically confirmed as TC (ICD-O-3 code: 8240/3), AC (ICD-O-3 code: 8249/3) or LCNEC (ICD-O-3 code: 8013/3). The study period was set from 2004 to 2015, as the TMN classification and Collaborative Stage information was available in the database since 2004. Besides, we reclassified the TNM stage according to the eighth edition of AJCC/UICC TNM classification because the TNM system had multiple versions in the SEER database and did not apply to all patients [26]. Furthermore, considering its strong invasion ability and unique pathological characteristics limiting the surgical options, SCLC was not included in the present study [27, 28]. Although LCNEC has been found to harbor subpopulations of tumors with SCLC, surgery should be offered to medically fit patients with both early and locally advanced LCNEC [29–31]. Therefore, we enrolled patients with LCNEC in this study. Patients were excluded who (1) aged 18 years; (2) had a diagnosis of any other cancer; (3) did not undergo radical surgery with systematic LN dissection; (4) had the diagnosis lacking pathological evidence; (5) were at pN0/pN3 disease; (6) had distant metastasis (M1); (7) received preoperative radiotherapy; (8) survived less or equal to 30 days after surgery; (9) had missing data on race, laterality, tumor location, radiotherapy, TNM staging system, and survival outcomes.

### 2.3 Variable extraction, preparation, grouping, and calculation

The baseline demographics data including age at diagnosis (≤62 and >62), sex (male and female), and race (white, black, and other) were extracted from the SEER database. Data on baseline tumor-related characteristics included primary site (upper lobe, middle lobe, lower lobe, and other), laterality (right and left), histotypes (TC, AC, and LCNEC), tumor differentiation (well/moderately differentiated, poorly differentiated /undifferentiated, and unknown), T classification (T1, T2, T3, and T4), and N classification (N1 and N2). In addition, treatment information including surgical intervention (sublobectomy, lobectomy, and pneumonectomy), radiotherapy (yes and no/unknown), chemotherapy (yes and no/unknown), NDLN, and NPLN was also extracted from the database. LODDS was calculated as: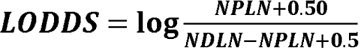. To avoid an infinite number, 0.50 was added to both the numerator and denominator. Lung cancer-specific survival (CSS) and overall survival (OS) were two of the study endpoints. The period from diagnosis to all-cause death was referred to as OS, while the time from diagnosis to LNET-related death was referred to as CSS. For censored data, the follow-up duration was computed as the number of months between diagnosis and death or the last follow-up (December 31, 2016).

### 2.4 Statistical analysis strategy

Baseline characteristics of study cohort stratified by LODDS were compared by using Student *t* test, Mann-Whitney test, Pearson’s χ^2^ test, or Fisher’s exact test as appropriate. Continuous variables were displayed with the mean (standard deviation [SD]) or the median (interquartile range [IQR]), while categorical variables were reported as count and percentages.

First, we analyzed LODDS as a continuous variable. A smooth curve was drawn to estimate the association between LODDS and its hazard ratios (HRs) using restricted cubic spline regression analysis according to the method of Chen et al [32]. Second, we took LODDS as a categorical variable. LODDS was dichotomized via the X-tile software [33]. Survival curves were calculated using the Kaplan-Meier method and compared using the log-rank test. To assess the connection between LODDS and each study outcome, a multivariable Cox proportional-hazards regression model was used, controlling for relevant confounders. These confounders were selected based on their associations with the study endpoints of interest or a change in effect estimate of over 10%. The relatively low LODDS group was used as a reference group, and the results are presented as HRs with 95% confidence intervals (CIs). Third, a series of subgroup analyses were performed according to age at diagnosis, sex, primary site, laterality, histotypes, tumor differentiation, T classification, and N classification.

All statistical analyses were performed using R software (version.3.6.14.1.0; The R Project for Statistical Computing, TX, USA; http://www.r-project.org) and EmpowerStats (version 2.0; http://www.empowerstats.com). Two-tailed *P* < 0.050 was deemed as statistical significance.

### 2.5 Construction and validation of nomograms

Patients with purchased/referred care delivery areas (PRCDA) of East, Northern plains, and Alaska were regarded as the derivation cohort, while the external validation cohort includes patients with PRCDA of Pacific coast and Southwest. StepAIC algorithm was used in multivariable Cox regression analysis to select the predictors for the final models [34]. Using these identified prognostic factors, we constructed two nomograms for predicting 1-, 3-, and 5-year CSS and OS in LNET patients.

Several indexes and methods were used to assess the accuracy of our nomograms. First, Harrell’s concordance index (c-index) was calculated to quantify the discrimination performance of the nomograms. Second, a calibration curve, which is a diagram presenting the association between predicted probabilities and actual survival rate at 1, 3, and 5 years, along with a bootstrapped sample of the study cohort, was used to assess the calibration. Third, a comparison between our nomogram and the conventional eighth edition of the TNM staging system was conducted by calculating integrated discrimination improvement (IDI) and net reclassification improvement (NRI) [35]. Z test was used to examine the difference. Fourth, the decision curve analysis (DCA) was performed to test the clinical usefulness of the nomograms and TNM classification. Finally, to facilitate researchers’ and clinicians’ usage of our model, we created two user-friendly webservers for our nomograms.

## 3. Results

### 3.1 Patients characteristics and cutoff value for LOODS

The SEER database collected 11,870 patients diagnosed with LNETs from January 2004 to December 2015. After employing the inclusion and exclusion criteria, 506 patients remained in the final study cohort. The selection process were summarized in ***Figure 1***. Based on the results calculated by the X-tile software, the optimal cutoff value of LODDS was set as -0.33. Therefore, the study cohort was dichotomized into the low LODDS group (-1.44≤LODDS<-0.33; n=334) and the high LODDS group (-0.33≤LODDS≤1.14; n=172). Baseline demographic and clinicopathological features of participants stratified by LODDS were listed in ***Table 1*.** The median number of NDLN was 12.00 (IQR: 8.00-17.00) in the low LODDS group and 5.00 (IQR: 3.00-9.00) in the high LODDS group, whereas the median number of NPLN was 1.00 (IQR: 1.00-2.00) in the low LODDS group and 2.00 (IQR: 1.00-4.00) in the high LODDS group. Compared with the low LODDS group (-1.44 ≤LODDS<-0.33), the patients included in the high LODDS group (-0.33 ≤ LODDS≤1.14) were more likely to be diagnosed in the N2 classification (*P* <0.001), have more positive lymph nodes (*P* < 0.001) and receive radiotherapy (*P*=0.045). No difference was observed in age at diagnosis, sex, race, laterality, tumor site, histotype, differentiation, T classification, surgery, and chemotherapy between the two cohorts (all *P-*values > 0.050).

**Figure 1.**
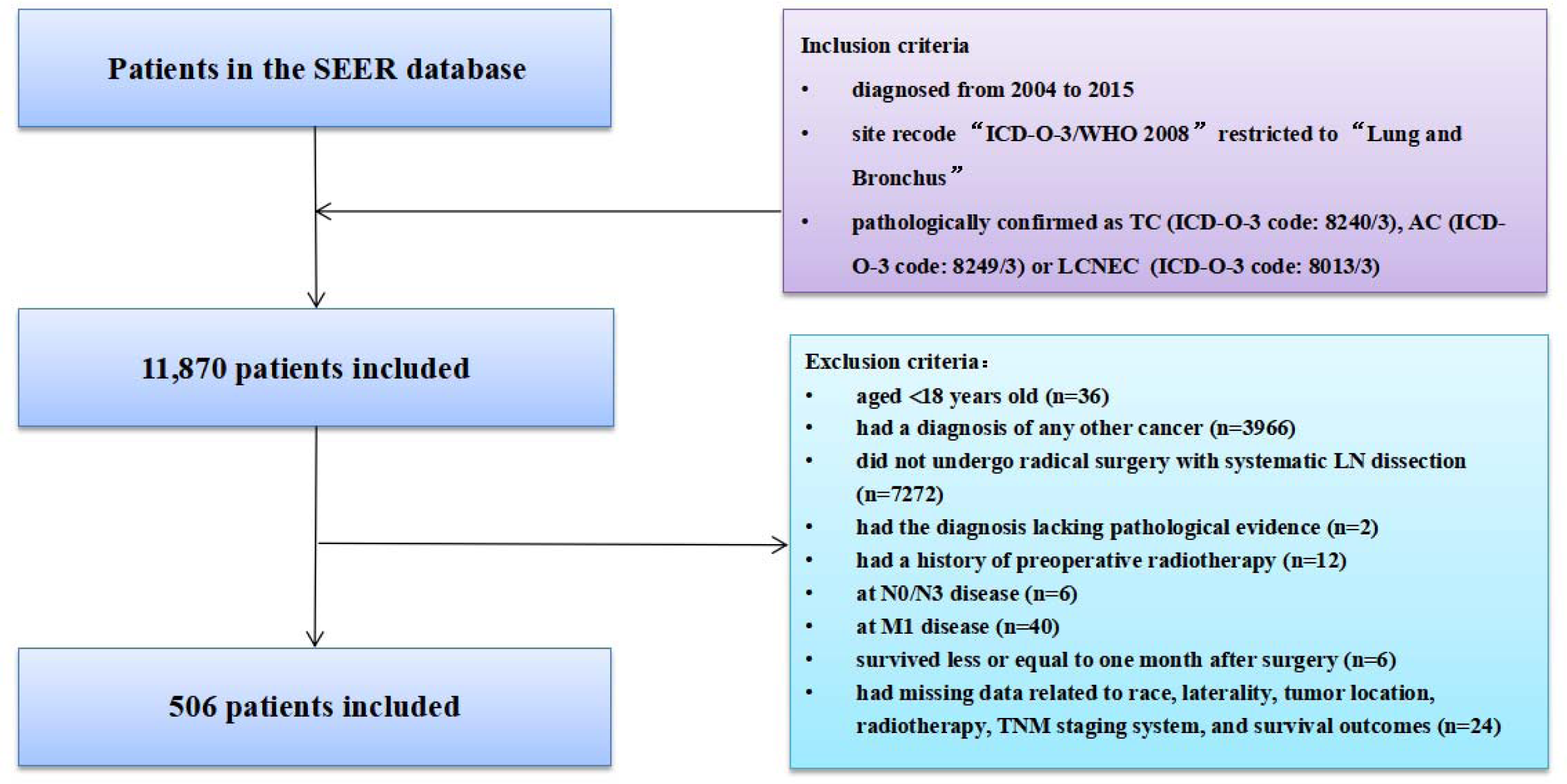
Selection of study cohort from the SEER database. AC: atypical carcinoid; LCNEC: large cell neuroendocrine carcinoma; SCLC: small cell lung carcinoma; SEER: Surveillance, Epidemiology, and End Results; LN: lymph node; TNM: tumor-node-metastasis.

**Table 1.**
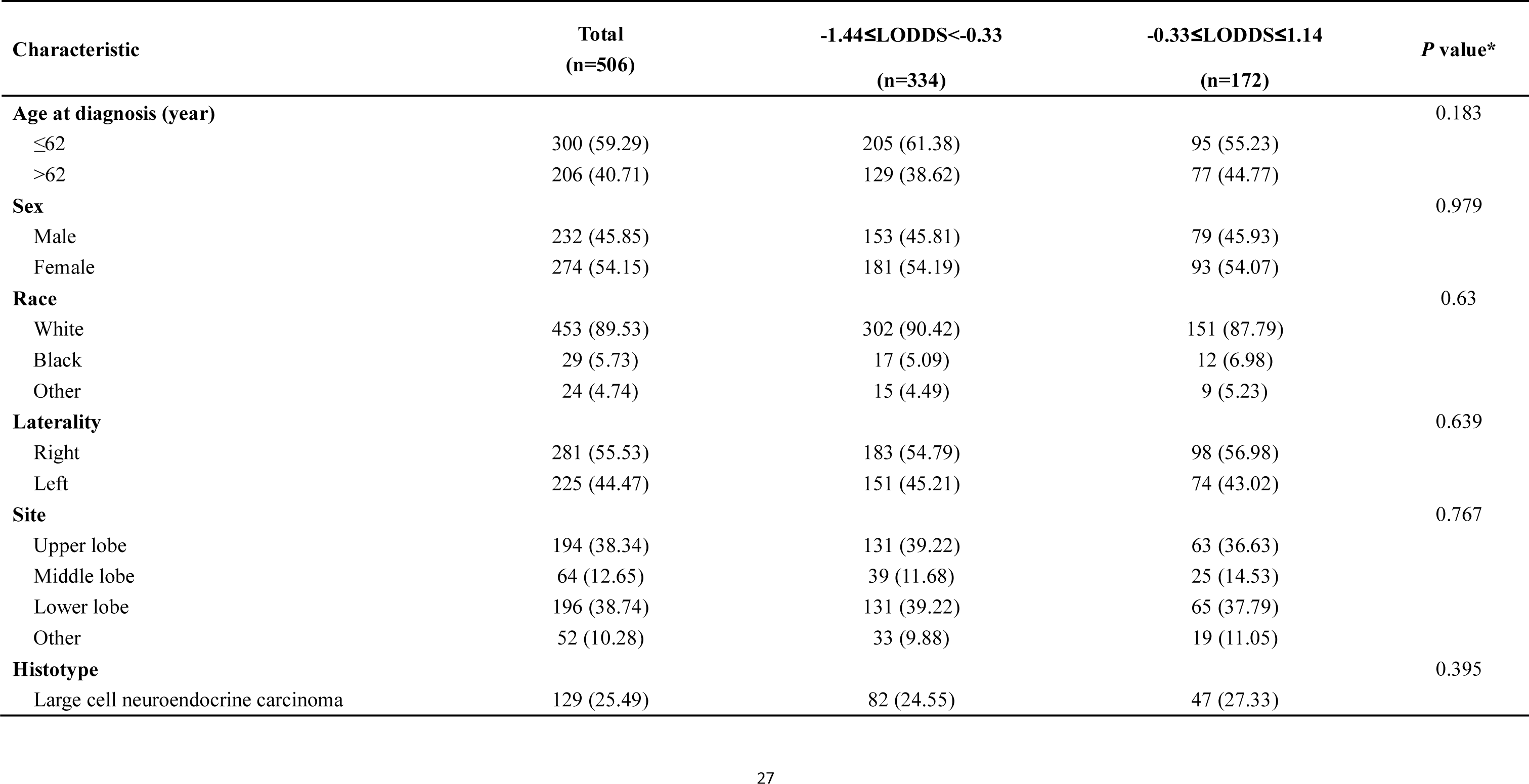
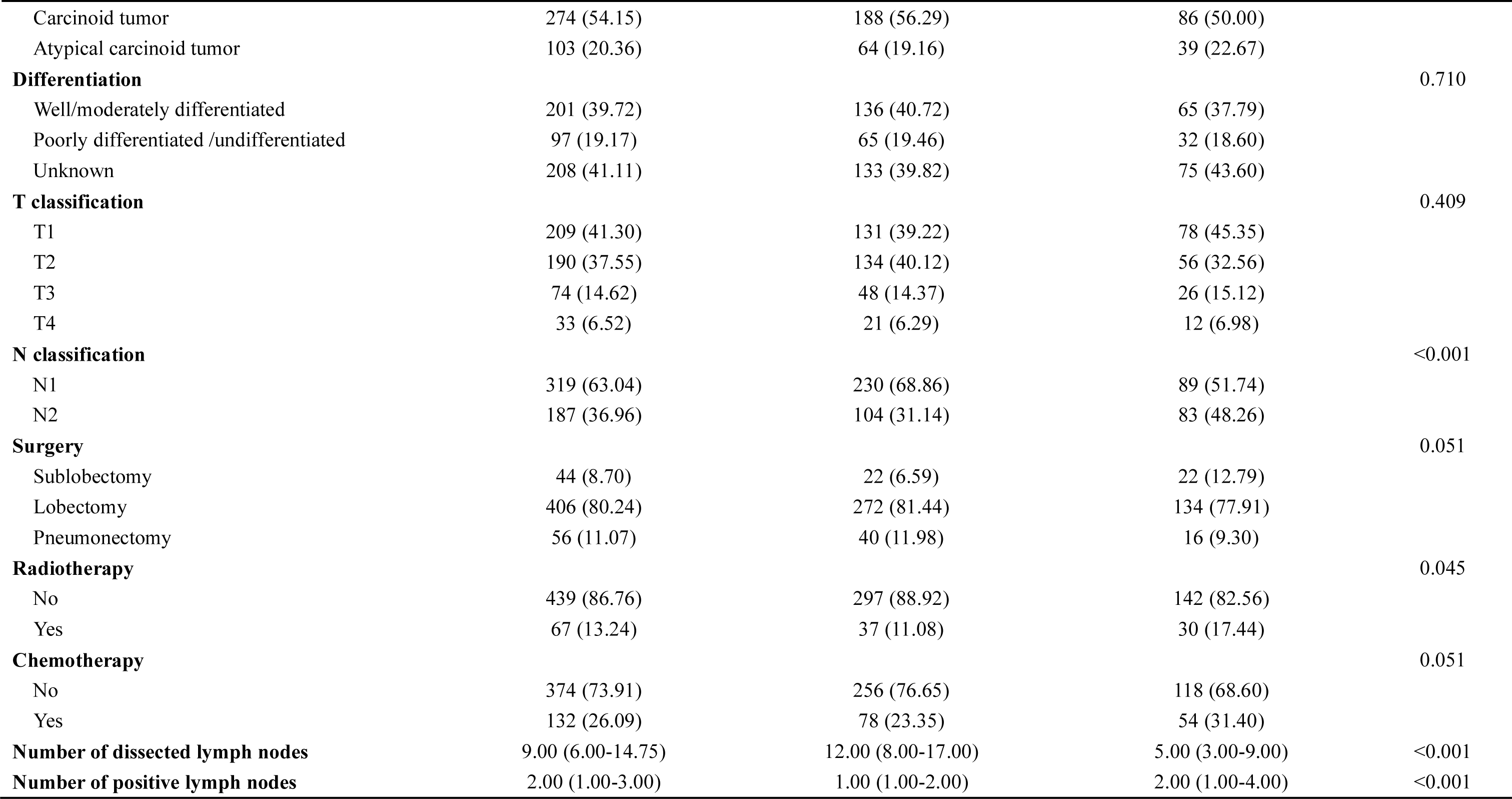
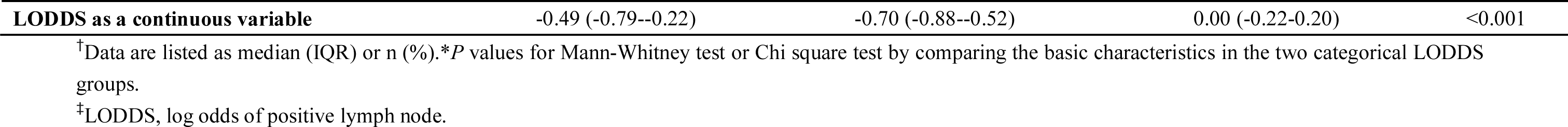
Baseline characteristics of the entire dataset stratified by LODDS

### 3.2 Restricted cubic spline analysis

We performed a common Cox regression analysis using LODDS as a continuous variable and found the adjusted HRs of LODDS for CSS and OS were 2.21 (95% CI: 1.52-3.20, *P*<0.001) and 1.51 (95%CI: 1.10-2.06, *P*=0.010). Second, two piece-wise Cox regression analyses were conducted and the inflection points were found (-0.27 for CSS, -0.26 for OS, ***Table S1***). For the LODDS > -0.27, every 1 increase in LODDS was associated with a 321% increase in CSS (*P*<0.001), while for the LODDS> -0.27, every 1 increase in LODDS was associated with a 24% increase in CSS (*P*=0.555). For a LODDS<-0.26, every 1 increase in LODDS was associated with a 135% increase in OS (*P*<0.001), while for a LODDS>-0.26, every 1 increase in LODDS was associated with a 4% decrease in OS (*P*=0.906). As shown in the restricted cubic spline analyses (***Figure 2***), the HRs for LODDS as a continuous variable tends to show a saturation effect.

**Figure 2.**
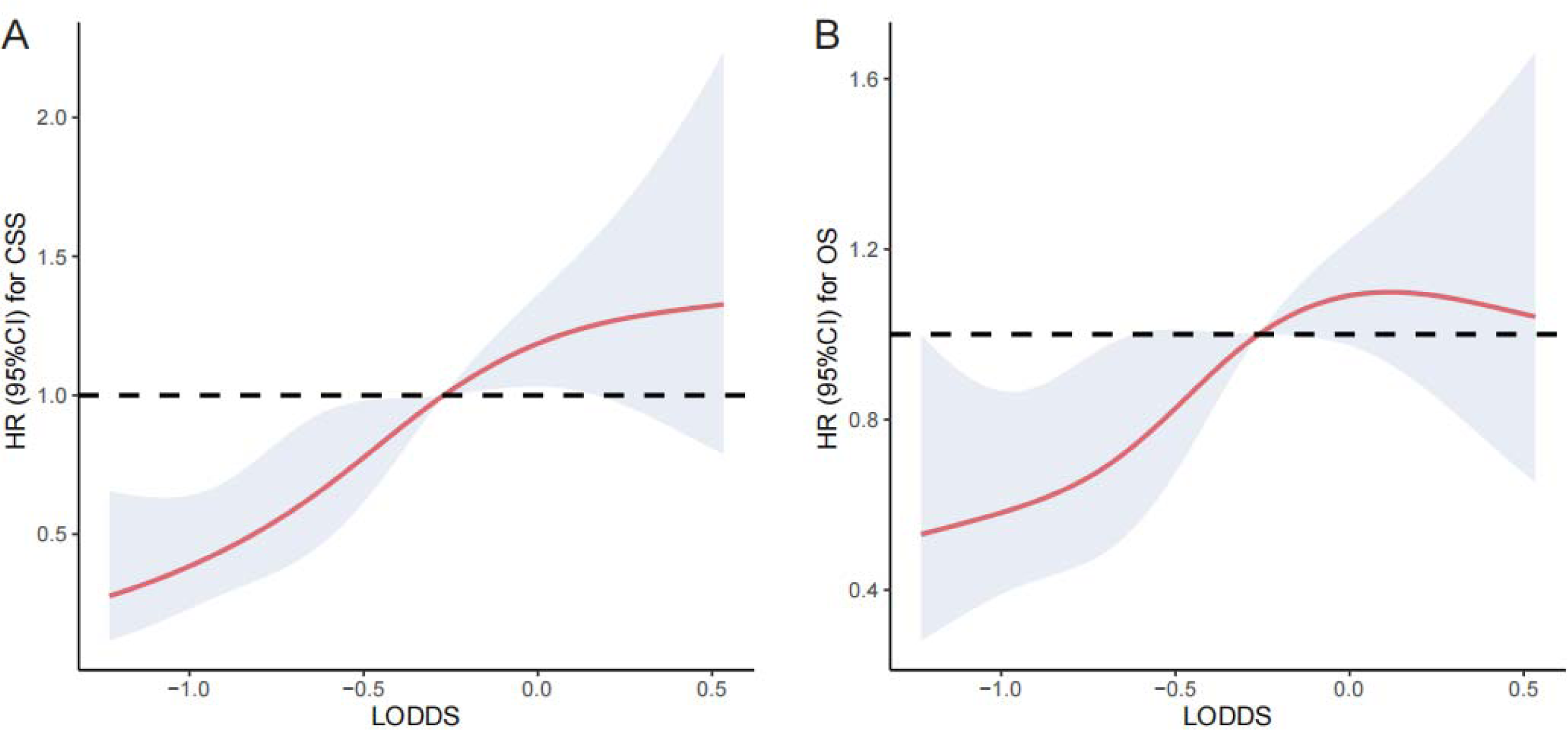
Restricted cubic spline fitting for the association between LODDS levels with the HRs of LODDS for CSS (A) and OS (B). LODDS: log odds of positive lymph nodes; CSS: lung cancer-specific survival; OS: overall survival; HR: hazard ratio; CI: confidence interval.

### 3.3 Survival analysis

The median follow-up time for the entire cohort was 42 months (IQR, 19-77 months). A total of 180 (35.57%) patients died from any cause, and 135 (26.68%) patients died from LNET-related death at the end of the study period. The cumulative 1-, 3-, and 5-year CSS and OS rates for patients stratified by LODDS were summarized in ***Tables S2***. The survival curves showed that patients with the higher values of LODDS (-0.33≤LODDS≤ 1.14) were associated with worse prognosis (log-rank test: *P*=0.001 for CSS; *P*=0.005 for OS, ***Figures S1***). Based on the entire cohort, the univariable Cox regression model suggested that age at diagnosis, race, site, histotype, differentiation, T classification, N classification, surgery, radiotherapy, chemotherapy, and LODDS were all significant variables for CSS in ***Table S3*.** The multivariable Cox regression analysis demonstrated that participants in the high LODDS group (-0.33≤LODDS≤1.14) was related to reducing survival compared to those in the low LODDS group (-1.44≤LODDS<-0.33) (HR=2.06, 95% CI: 1.43-2.97, *P*<0.001) (***Table 2 and Table S3***). The multivariable Cox regression analysis for OS yielded similar results (HR=1.66, 95% CI: 1.22-2.26, *P*=0.001, ***Table 2 and Table S4***). To eliminate the potential bias caused by the LN fragments, we performed a sensitivity analysis by restricting the resected LN count to fewer than or equal to 20, and found that LODDS remained statistically significant (CSS: HR=2.16, 95% CI: 1.46-3.19, *P*<0.001; OS: HR=1.75, 95% CI: 1.25-2.43, *P*=0.001) (***Table 2 and Table S5-6***). Furthermore, the extent of LN management should be in accordance with the IASLC recommendations, which involves a minimum of 6 nodes/stations, therefore we excluded patients with the examined LN count to fewer than 6. The multivariable Cox regression analysis yielded similar results (CSS: HR=2.84, 95% CI: 1.78-4.50, *P*<0.001; OS: HR=2.37, 95% CI: 1.59-3.54, *P*<0.001) (***Table 2 and Table S7-8***).

**Table 2.** HRs for multivariable analysis

| Cohort | CSS |  | OS |  |
| --- | --- | --- | --- | --- |
|  | HR (95%CI) | *P* value | HR (95%CI) | *P* value |
| **Entire dataset** |  |  |  |  |
| Univariable analysis | 1.73 (1.23-2.43) | 0.002 | 1.52 (1.13-2.04) | 0.006 |
| Multivariable analysis | 2.06 (1.43-2.97) | <0.001 | 1.66 (1.22-2.26) | 0.001 |
| **Patients with NDLN≤20** |  |  |  |  |
| Univariable analysis | 1.89 (1.32-2.72) | 0.001 | 1.65 (1.20-2.26) | 0.002 |
| Multivariable analysis | 2.16 (1.46-3.19) | <0.001 | 1.75 (1.25-2.43) | 0.001 |
| **Patients with NDLN≥6** |  |  |  |  |
| Univariable analysis | 2.16 (1.42-3.28) | <0.001 | 1.97 (1.37-2.84) | <0.001 |
| Multivariable analysis | 2.84 (1.78-4.50) | <0.001 | 2.37 (1.59-3.54) | <0.001 |

<sup>†</sup>CSS, cancer-specific survival; OS, overall survival; HR, hazard ratio; CI, confidence interval; NDLN, number of dissected lymph node.

### 3.4 Subgroup analysis

For CSS, subgroup analyses showed the higher LODDS (-0.33 ≤ LODDS≤1.14) was also associated with deteriorative mortality in most strata (***Table S9***). Even among patients with the same N stage, LODDS still could distinguish and stratify patients into different risk groups. (N1 stage subgroup: HR=1.99, 95% CI: 1.14-3.45, *P*=0.015; N1 stage subgroup: HR=2.24, 95% CI: 1.27-3.96, *P*=0.006). For OS, in the N1 stage subgroup, the HR remained the trend of increase despite the results were not statistically significant (HR=1.56, 95% CI: 0.98-2.49, *P*=0.060).

### 3.5 Construction and validation of the nomogram

Prognostic nomograms for CSS and OS were established including significant indicators selected by StepAIC algorithm in multivariable Cox regression analysis. For nomogram construction and validation, among the final study cohort including 506 patients, 300 of them were assigned to the derivation cohort (PRCDA=East, Northern plains, and Alaska) and 206 of them were assigned to the validation cohort (PRCDA=Pacific coast, Southwest). The comparison of baseline characteristics between the derivation cohort and validation cohort was summarized in ***Table S10*.** No difference was observed in most variables except sex. The nomogram of CSS showed that histotype contributed the most to the prognosis, followed by age at diagnosis, LODDS, surgery, and chemotherapy (***Fig 3*** and ***Table 3***). Similarly, the nomogram of OS showed that histotype contributed the most to the prognosis, followed by age at diagnosis, chemotherapy, LODDS, and radiotherapy (***Fig 3*** and ***Table 4***). The top point reference scale of the nomograms assigned a score for each category of these predictive variables. After adding up the total score and locating the sum on the total points reference scale, a straight line was drawn to the bottom survival probability scale to find the estimated 1-/3-/5-survival rate.

**Figure 3.**
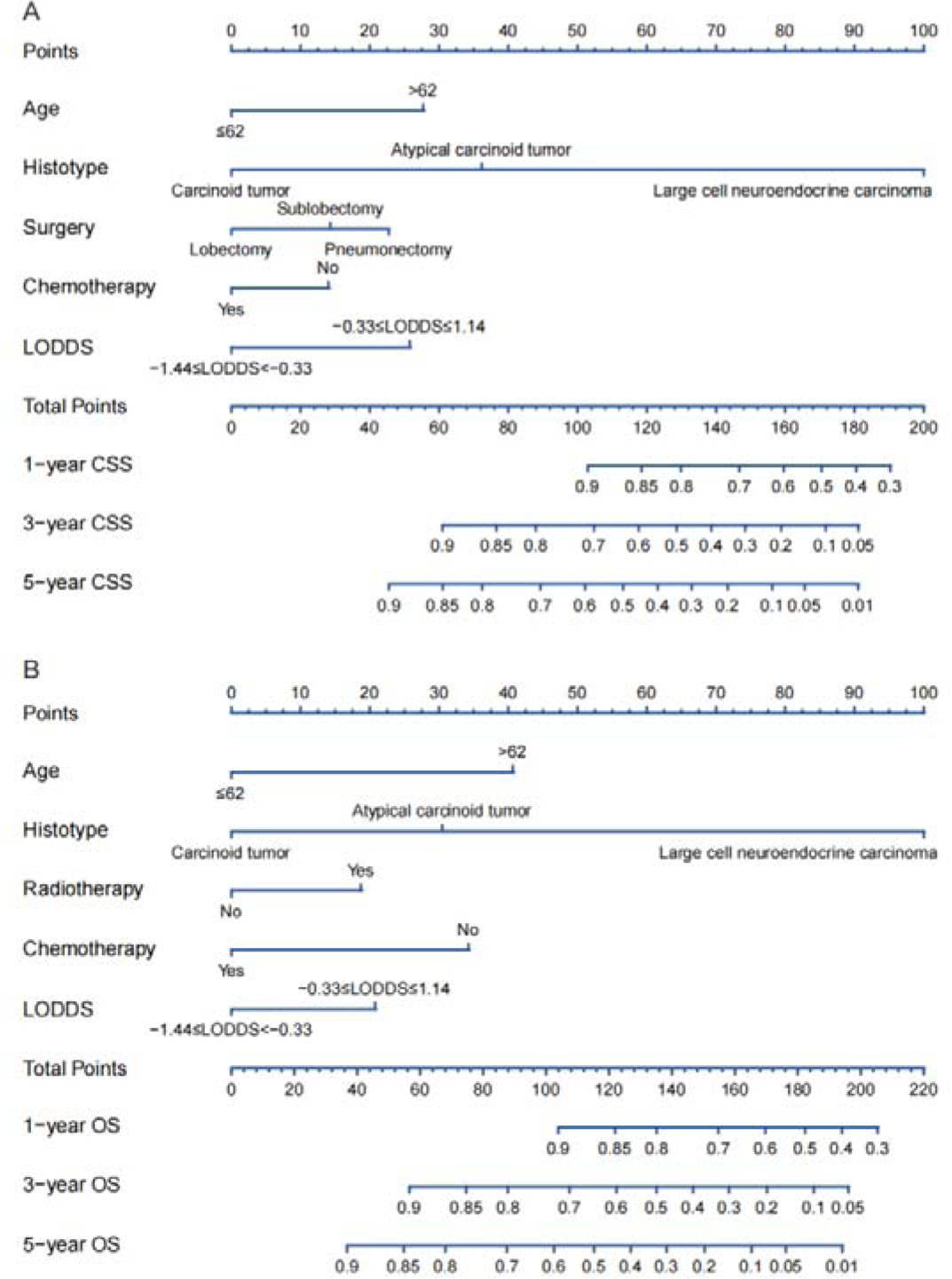
Nomograms to predict 1-, 3- and 5-year CSS (A) and OS (B) for patients with node-positive lung neuroendocrine carcinoma after surgery. CSS: lung cancer-specific survival; OS: overall survival; LODDS: log odds of positive lymph nodes.

**Table 3.**
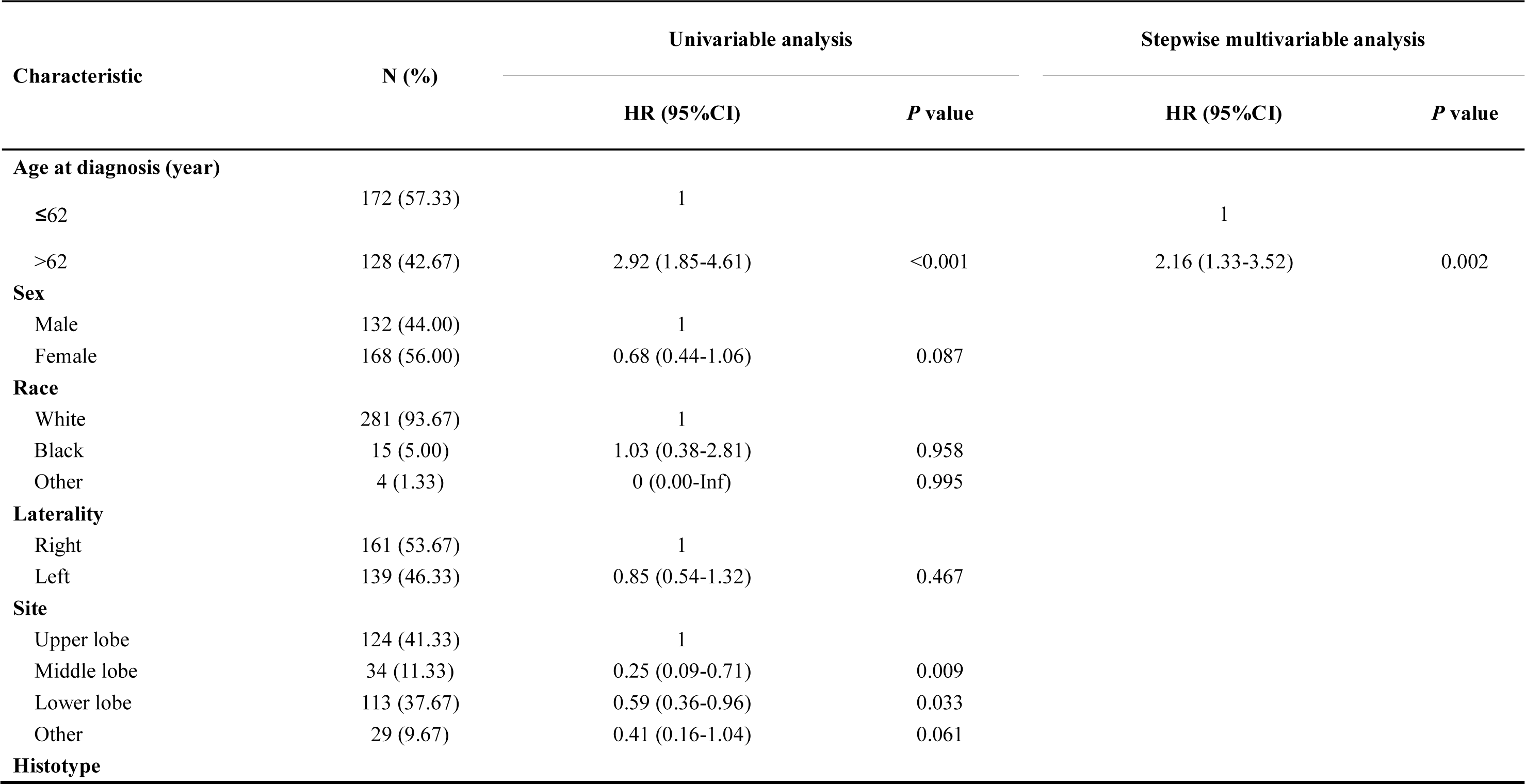
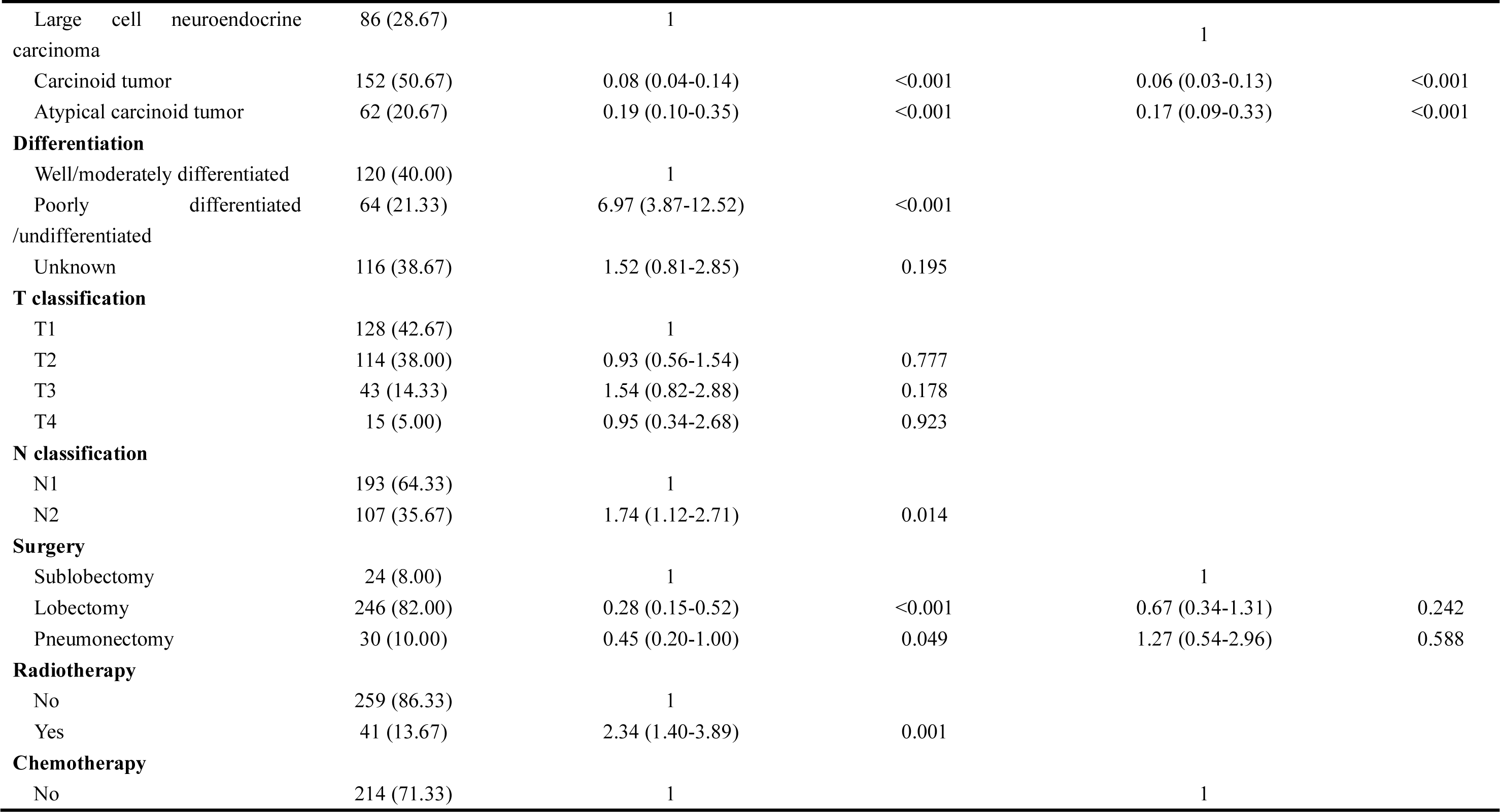
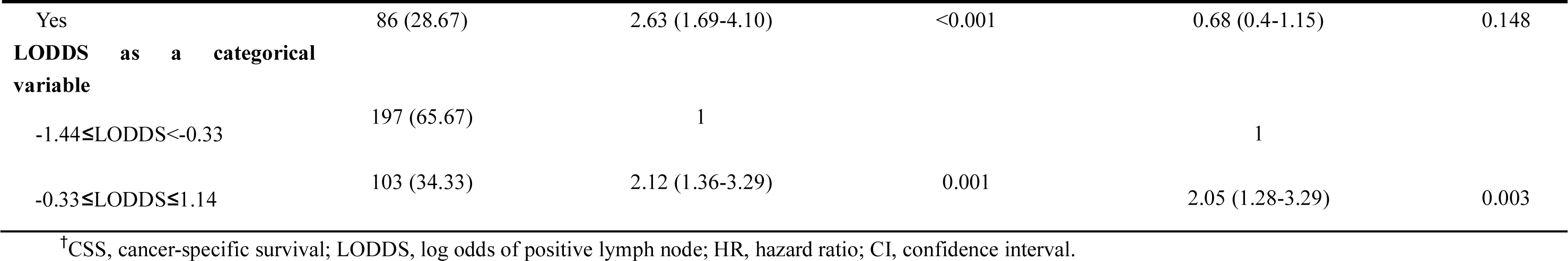
Univariable and stepwise multivariable Cox proportional regression analysis for CSS of the derivation dataset

**Table 4.** Univariable and stepwise multivariable Cox proportional regression analysis for OS of the derivation dataset

| Characteristic | N (%) | Univariable analysis |  | Stepwise multivariable analysis |  |
| --- | --- | --- | --- | --- | --- |
|  |  | HR (95%CI) | *P* value | HR (95%CI) | *P* value |
| **Age at diagnosis (year)** |  |  |  |  |  |
| ≤62 | 172 (57.33) | 1 |  | 1 |  |
| >62 | 128 (42.67) | 3.5 (2.36-5.20) | <0.001 | 2.65 (1.75-4.01) | <0.001 |
| **Sex** |  |  |  |  |  |
| Male | 132 (44.00) | 1 |  |  |  |
| Female | 168 (56.00) | 0.62 (0.43-0.91) | 0.014 |  |  |
| **Race** |  |  |  |  |  |
| White | 281 (93.67) | 1 |  |  |  |
| Black | 15 (5.00) | 1.11 (0.49-2.53) | 0.801 |  |  |
| Other | 4 (1.33) | 0 (0.00-Inf) | 0.994 |  |  |
| **Laterality** |  |  |  |  |  |
| Right | 161 (53.67) | 1 |  |  |  |
| Left | 139 (46.33) | 0.87 (0.59-1.26) | 0.453 |  |  |
| **Site** |  |  |  |  |  |
| Upper lobe | 124 (41.33) | 1 |  |  |  |
| Middle lobe | 34 (11.33) | 0.41 (0.20-0.87) | 0.020 |  |  |
| Lower lobe | 113 (37.67) | 0.74 (0.49-1.12) | 0.149 |  |  |
| Other | 29 (9.67) | 0.69 (0.35-1.36) | 0.284 |  |  |
| **Histotype** |  |  |  |  |  |
| Large cell neuroendocrine carcinoma | 86 (28.67) | 1 |  |  |  |
| Carcinoid tumor | 152 (50.67) | 0.13 (0.08-0.20) | <0.001 | 0.09 (0.05-0.16) | <0.001 |
| Atypical carcinoid tumor | 62 (20.67) | 0.23 (0.13-0.39) | <0.001 | 0.19 (0.11-0.34) | <0.001 |
| **Differentiation** |  |  |  |  |  |
| Well/moderately differentiated | 120 (40.00) | 1 |  |  |  |
| Poorly differentiated/undifferentiated | 64 (21.33) | 6.22 (3.70-10.46) | <0.001 |  |  |
| Unknown | 116 (38.67) | 1.98 (1.18-3.34) | 0.010 |  |  |
| **T classification** |  |  |  |  |  |
| T1 | 128 (42.67) | 1 |  |  |  |
| T2 | 114 (38.00) | 1.12 (0.74-1.70) | 0.586 |  |  |
| T3 | 43 (14.33) | 1.35 (0.76-2.40) | 0.301 |  |  |
| T4 | 15 (5.00) | 0.91 (0.36-2.31) | 0.849 |  |  |
| **N classification** |  |  |  |  |  |
| N1 | 193 (64.33) | 1 |  |  |  |
| N2 | 107 (35.67) | 1.76 (1.21-2.57) | 0.003 |  |  |
| **Surgery** |  |  |  |  |  |
| Sublobectomy | 24 (8.00) | 1 |  |  |  |
| Lobectomy | 246 (82.00) | 0.38 (0.21-0.67) | 0.001 |  |  |
| Pneumonectomy | 30 (10.00) | 0.49 (0.23-1.03) | 0.058 |  |  |
| **Radiotherapy** |  |  |  |  |  |
| No | 259 (86.33) | 1 |  | 1 |  |
| Yes | 41 (13.67) | 1.98 (1.26-3.12) | 0.003 | 1.57 (0.9-2.72) | 0.110 |
| **Chemotherapy** |  |  |  |  |  |
| No | 214 (71.33) | 1 |  | 1 |  |
| Yes | 86 (28.67) | 1.86 (1.26-2.73) | 0.002 | 0.44 (0.26-0.75) | 0.003 |
| **LODDS as a categorical variable** |  |  |  |  |  |
| -1.44≤LODDS<-0.33 | 197 (65.67) | 1 |  | 1 |  |
| -0.33≤LODDS≤1.14 | 103 (34.33) | 1.68 (1.15-2.45) | 0.007 | 1.65 (1.12-2.42) | 0.011 |

<sup>†</sup>LODDS, log odds of positive lymph node; OS, overall survival; HR, hazard ratio; CI, confidence interval.

The nomogram was validated internally in the derivation cohort and externally in the validation cohort. C-indexes of CSS for derivation cohort and validation cohort were 0.845 (0.804-0.886), and 0.821 (0.771-0.871). C-indexes of OS for derivation cohort and validation cohort were 0.821 (0.783-0.858), and 0.811 (0.766-0.856). Additionally, the calibration plots (***Fig 4***) revealed that the points were close to the 45-degree line, indicating a good similarity between the nomogram-predicted and actual survival states.

**Figure 4.**
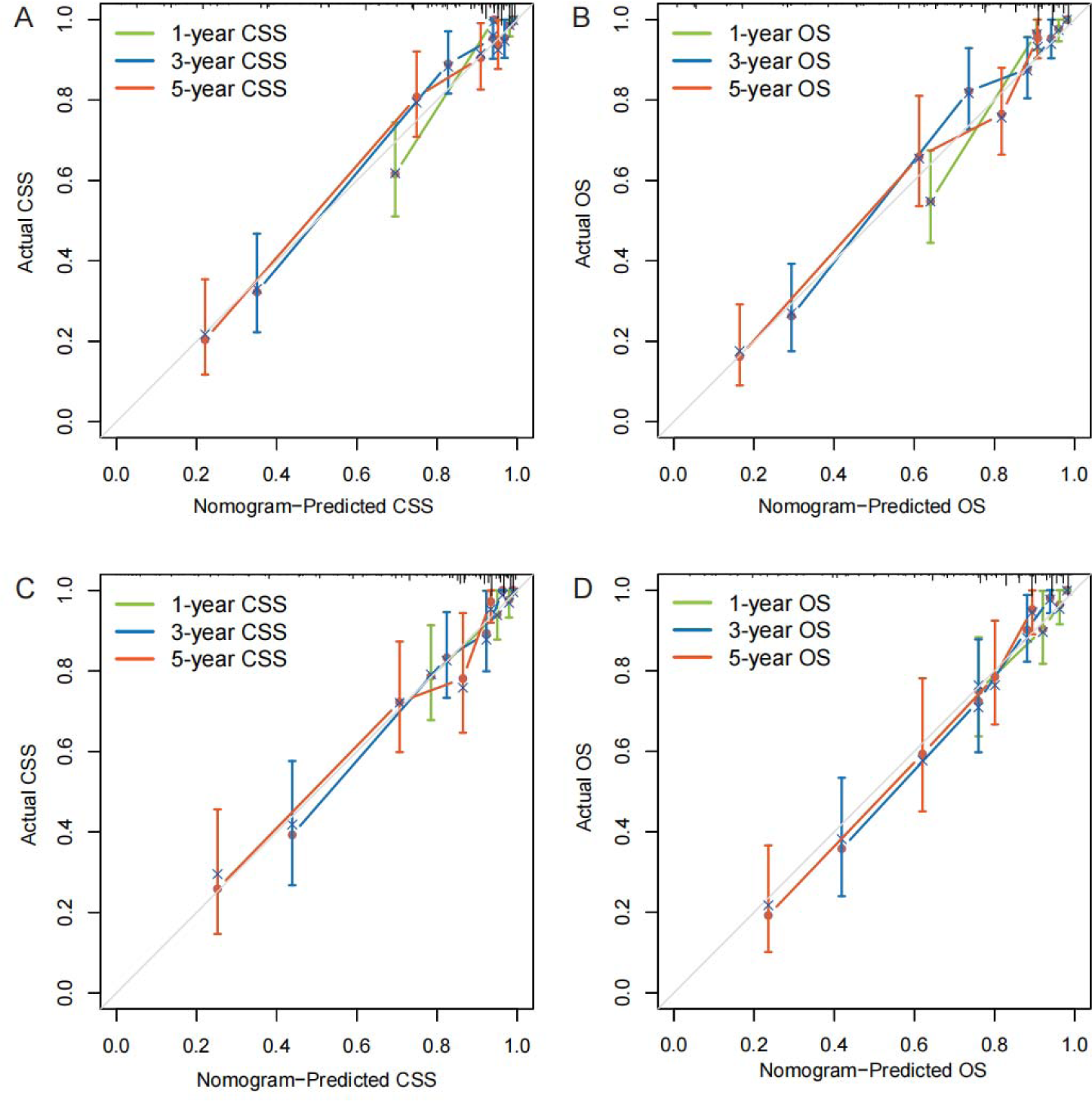
Calibration plots of the nomograms to predict CSS and OS of the derivation dataset (A, B) and external validation dataset (C, D). CSS: lung cancer-specific survival; OS: overall survival.

### 3.6 Comparison of the nomogram and the eighth edition TNM staging system

The comparisons between the nomograms and the TNM staging system were also performed (***Table S11***). Analysis of accuracy showed that the NRI for the 1-, 3-, and 5-year CSS were 0.721 (95% CI: 0.825-0.554, *P*<0.001), 0.645 (95% CI: 0.743-0.414, *P*<0.001) and 0.627 (95% CI: 0.768-0.403, *P*<0.001) in the validation cohorts, respectively. Similarly, in the validation set, the IDI for 1-, 3-, and 5-years CSS were 0.249 (95% CI: 0.389-0.161, *P*<0.001), 0.391 (95% CI: 0.519-0.243, *P*<0.001), and 0.42 (95% CI: 0.537-0.265, *P*<0.001), respectively. Furthermore, the DCA curve was applied to determine the clinical applicability of the nomogram and TNM staging system. The DCA curves showed that our nomograms were better than the TNM staging system, as it added more net benefits compared with the TNM classification for almost all threshold probabilities based on the derivation cohort and validation cohort (***Fig S2 and S3***).

### 3.7 Development of webservers for convenient clinical use

Two dynamic, visualized and publicly accessible online nomograms based on the multivariable Cox regression models were built (CSS: https://drboidedwater.shinyapps.io/DynNom-CSS-lungneuroendocrinecarcinoma/; OS: https://drboidedwater.shinyapps.io/DynNom-OS-lungneuroendocrinecarcinoma/) (***Fig S4***). The webservers may generate estimated survival probability and Kaplan-Meier curves by entering the covariates.

## 4. Discussion

LNETs constitute a unique clinical subgroup of primary pulmonary tumors. Due to their relatively low incidence, no specific staging system exists for LNETs. An exact and reasonable assessment of the lymph node status plays a crucial role in the staging and prognosis evaluation of patients with LNCTs. In this study, we found that LODDS was independently associated with long-term clinical outcomes among patients with resectable LNETs. These results were robust to restricted cubic spline analysis and a series of sensitivity analyses. Second, we constructed a visualized and publicly accessible online nomogram, incorporating LODDS and routinely available demographic, staging and treatment information, to predict the survival probability for individual LNET patients. To our knowledge, the present study is the first to explore the prognostic value of LODDS for LNET based on a multi-center cohort with a relatively large sample size.

The involvement of regional lymph nodes in malignancies has been considered as one of the most important prognostic factors. The eighth edition of the AJCC/UICC staging system of NSCLC classified metastasis to ipsilateral peribronchial and/or hilar nodes and intrapulmonary nodes as N1 classification, and metastasis into ipsilateral mediastinal and/or subcarinal nodes into N2 classification, without taking into account numbers of examined and metastatic lymph nodes [36]. LODDS is a new LN ratio-based index and has been identified as an independent predictor in many malignancies such as rectal cancer [37], pancreatic cancer [38], gallbladder cancer [39], gastric cancer [40], and colon cancer [41]. Recently, several studies were attempting to explore the prognostic value of LODDS for NSCLC. In 2020, we did a research and found that the high value of LODDS>-0.37 was independently associated with worse survival in patients with node-positive lung squamous cell carcinoma [20]. Dziedzic et al. [19] found that it is possible to discriminate NSCLC patients more effectively by using LODDS compared to conventional N classification. Deng et al. [18] found LODDS and lymph node ratio (LNR) staging schemes outperformed those of NPLN for predicting OS and CSS among patients with node-positive NSCLC. However, most previous studies only focused on NSCLC, and few reports have explored the prognostic value of LODDS in LNETs.

In the current study, either as a continuous or as a categorical variable, the high value of LODDS was associated with worse survival for N1/N2 stage patients with LNETs. However, LODDS must be used and calculated with caution, because the value of LODDS is influenced by the number of dissected LN. Therefore, a series of sensitivity analyses were performed. For LNETs, the extent of LN management should conform to the International Association for the Study of Lung Cancer (IASLC) recommendations, which involves a minimum of 6 nodes/stations, 3 of which should be mediastinal including the subcarinal station [42]. Considering that the accurate value of LODDS was dependent on the adequate NDLN, we excluded patients with examined LN count less than 6 and found that LODDS could still serve as an independent predictor for LNETs. In addition, it is quite easy to damage the integrity of the LN during surgery, especially when the LNs are adhesive to each other or difficult to be separated from the dissected tissues [43]. To avoid the potential bias led by fragmented LNs, we excluded patients with examined LN count of more than 20, and the results did not change. Furthermore, in the subgroup stratified by different N stages (N1 and N2 stages), LODDS could still discriminate the high-risk population from the low-risk one. Additionally, to our surprise, N classification was excluded from our nomograms. The possible reason is that the anatomical definition of LN station is more complicated and dependent on surgery quality, which thus might potentially cause misclassification of the stage, and thus be replaced by LODDS [44, 45]. In summary, LODDS is the ratio-based LN staging system that combines NPLN and NNLN, which might be superior to some number-based LN assessment methods. Furthermore, in LODDS, the numerator and denominator are both added with a value of 0.5, eliminating the singularities caused by null data, therefore LODDS might be used to estimate survival of node-negative patients, as opposed to LNR.

The LODDS was not the only prognostic factor included in our nomogram. Similar to previous studies [46–48], age at diagnosis, histotype, surgery, radiotherapy, and chemotherapy were independent prognostic factors of CSS or OS. In this study, the nomogram showed that histotype contributed the most to the prognosis, which indicated tumor histotype is a crucial determinant of the clinical behavior of LNETs. Our nomogram indicated that LCNEC showed the worst prognosis followed by AC and TC. Growing evidence also suggests that high-grade LCNEC is biologically distinct from low-grade TC and intermediate-grade AC in view of clinical behavior, pathologic features, molecular alterations as well as possible precursor lesions [49]. All-stage 5-year OS for LCNEC fluctuated between 13% and 57% [50]. Different from LCNEC, TC and AC are more commonly found in younger patients without smoking histories. AC is significantly more aggressive than TC, with a higher frequency of nodal and distant metastases, and 5-year survival of 60%. In this study, we did not include SCLC patients, because a unique feature of SCLC is that it is generally considered a nonsurgical disease [51].

The optimal type of surgical treatment for LNETs is controversial. The surgical approach depends on tumor size, location, and preoperative biopsy specimen assessment. Several studies reported that wedge resection may increase the risk of tumor recurrences, especially in node-positive TC or AC [52, 53]. Lobectomy is reported as superior to segmentectomy in terms of OS in some, but not all in pulmonary carcinoids [54–56]. Similar to these studies, our study showed that lobectomy was superior to sublobectomy and pneumonectomy in the nomogram for CSS. Furthermore, there is an absence of high-quality evidence to show whether or not chemotherapy could provide clinical benefits for LNETs. Although our nomogram showed that chemotherapy might be associated with more favorable prognoses. However, the results need to be interpreted with caution. In the SEER database, patients without receiving chemotherapy and those with unknown information about adjuvant therapy were classified into one category, which might lead to potential bias. Until now, for pulmonary carcinoids, routine adjuvant therapy may only be considered in selected fit patients (AC, N2 stage) with a particularly high risk of relapse [57].

Besides, Iyoda et al. [58] suggested that platinum-based adjuvant chemotherapy after surgery could help patients with LCNEC prevent recurrence. Because LNET is a heterogeneous disease, each LNET patient requires an individualized and timely risk assessment, which allows for more precise therapeutic strategies and medical resource allocation decisions. In this study, we developed and validated a nomogram to predict prognosis in patients with LNET. Our nomograms based on LODDS were more accurate and obtained more clinical net benefit than the conventional AJCC/UICC TNM staging system. In summary, the online nomograms, composed of several easily obtained predictors, could be a simpler way to engage clinicians in death risks, patient counseling, and decision-making. To put it another way, LNET patients with poorer clinical results estimated by nomograms may require more aggressive therapy. [28].

Several limitations of this study should be noted. First, the SEER database lacks some detailed data, such as smoking history, some promising molecular markers (e.g. Ki-67), imaging techniques used before surgery, histological and morphological data (e.g. mitotic rate), type of resection (R0, R1, or R2), and use of systemic therapies. Therefore they could not be included as covariables in the multivariable Cox models. Second, information about postoperative comorbidities, and tumor recurrence is also not available in the database. Third, although we reclassified the TNM classification according to the eighth edition of AJCC/UICC TNM classification, the TNM staging system, which was derived from the SEER database’s collaborative stage system, is a combination of clinical and pathologic stages. Because of the distinction between clinical and pathologic stages, more subgroup analysis is required when using a single clinical or pathologic staging system.

## 5. Conclusions

LODDS was found to be useful in predicting survival outcomes in LNET patients who underwent surgery. Online dynamic nomograms including LODDS to evaluate CSS and OS were constructed. The well-executed nomograms may aid clinicians in providing reasonable, customized therapeutic strategies for LNET patients.

## CRediT authorship contribution statement

Suyu Wang and Yue Yu: Conceptualization, Methodology, Software, Investigation, Writing-original draft, Writing-review & editing. Juan Wei: Software, Data curation, Investigation, Writing-original draft. Yibin Guo and Qiumeng Xu: Visualization, Investigation. Xin Lv: Software, Validation. Meiyun Liu: Funding acquisition, Writing-review & editing.

## Data Availability

All data produced in the present study are available upon reasonable request to the authors

## Abbreviations

LNET: lung neuroendocrine tumor
TC: typical carcinoid
AC: atypical carcinoid
LCNEC: large cell neuroendocrine carcinoma
SCLC: small cell lung carcinoma
NSCLC: non-small cell lung cancer
AJCC: American Joint Committee on Cancer
UICC: Union for International Cancer Control
TNM: tumor-node-metastasis
LN: lymph node
NPLN: number of positive lymph node
LODDS: log odds of positive lymph nodes
SEER: Surveillance, Epidemiology, and End Results
NDLN: number of dissected lymph node
CSS: lung cancer-specific survival
OS: overall survival
SD: standard deviation
IQR: interquartile range
HR: hazard ratio
CI: confidence interval
PRCDA: purchased/referred care delivery area
c-index: concordance index
IDI: integrated discrimination improvement
NRI: net reclassification improvement
DCA: decision curve analysis
LNR: lymph node ratio
IASLC: International Association for the Study of Lung Cancer

## Tables S1-11

**Table S1.**
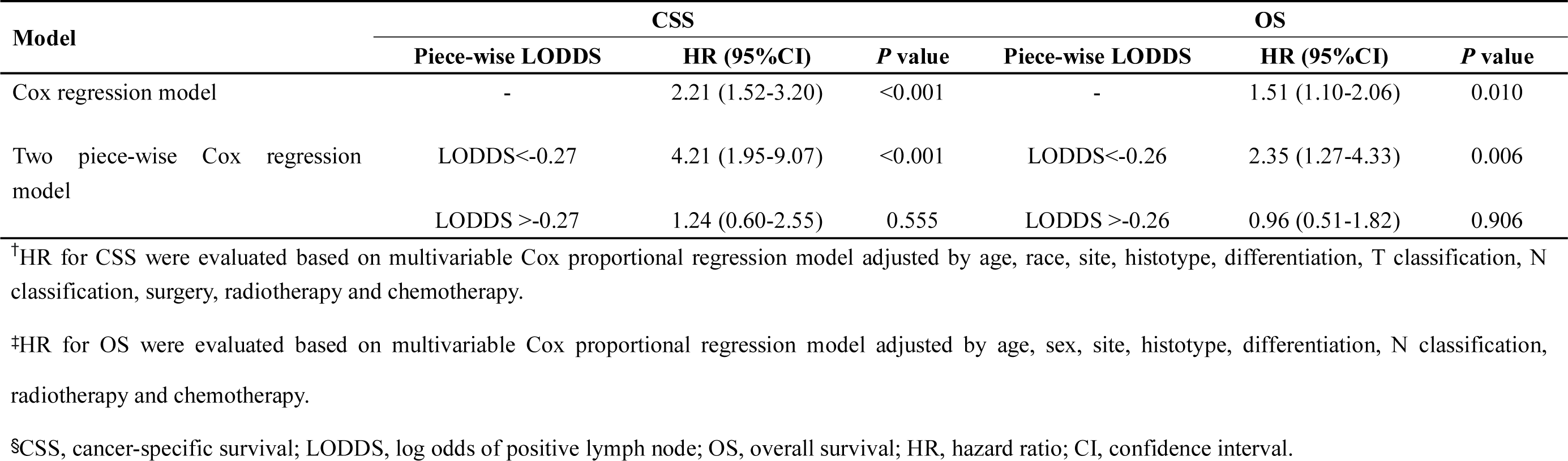
HR for LODDS as a continuous variable Model.

**Table S2.** 1-, 3-, and 5-year CSS and OS of different LODDS subgroups.

| Cohort | -1.44≤LODDS<-0.33 | -0.33≤LODDS≤1.14 |
| --- | --- | --- |
| 1-year CSS (%) | 93.83 (91.25-96.49) | 87.52 (82.66-92.66) |
| 3-year CSS (%) | 83.92 (79.82-88.23) | 70.57 (63.69-78.19) |
| 5-year CSS (%) | 76.83 (71.83-82.19) | 62.09 (54.37-70.9) |
| 1-year OS (%) | 90.96 (87.93-94.1) | 85.4 (80.28-90.86) |
| 3-year OS (%) | 78.7 (74.23-83.44) | 66.22 (59.19-74.08) |
| 5-year OS (%) | 70.21 (64.94-75.91) | 54.2 (46.46-63.23) |

<sup>†</sup>CSS, cancer-specific survival; LODDS, log odds of positive lymph node; OS, overall survival.

**Table S3.**
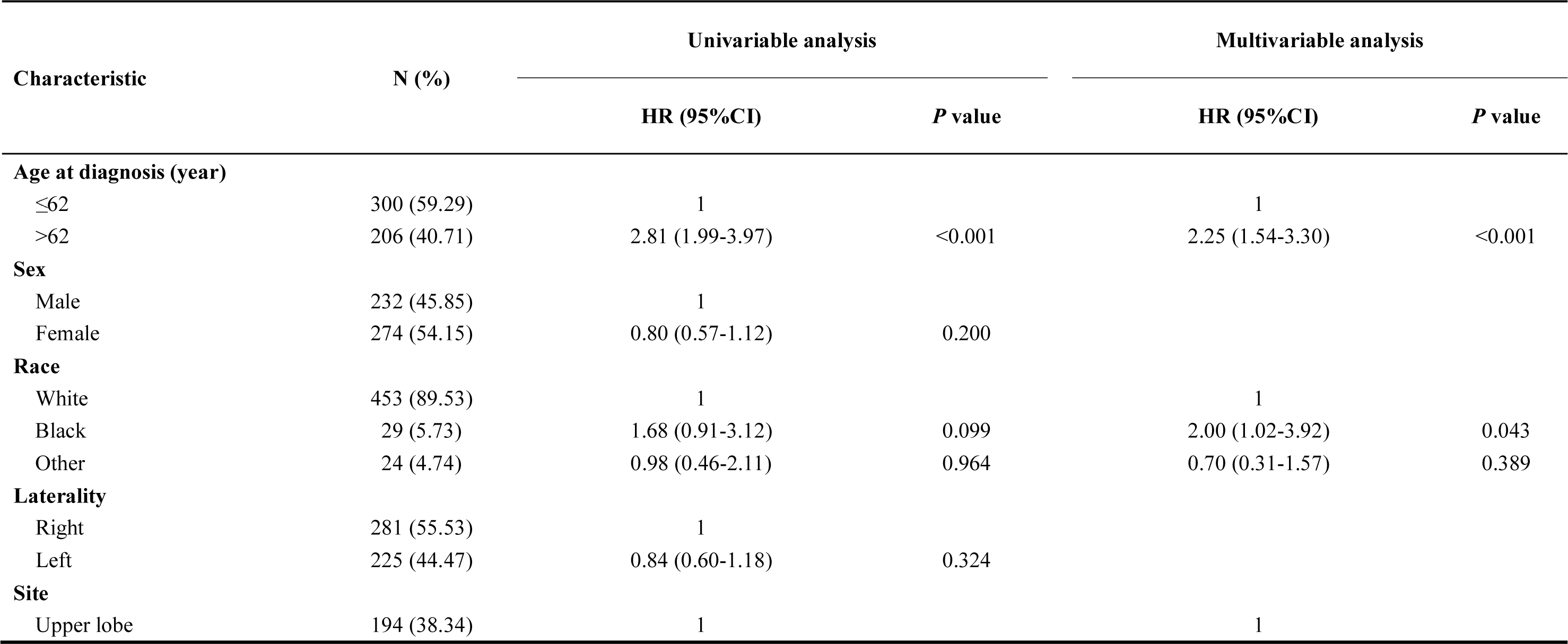
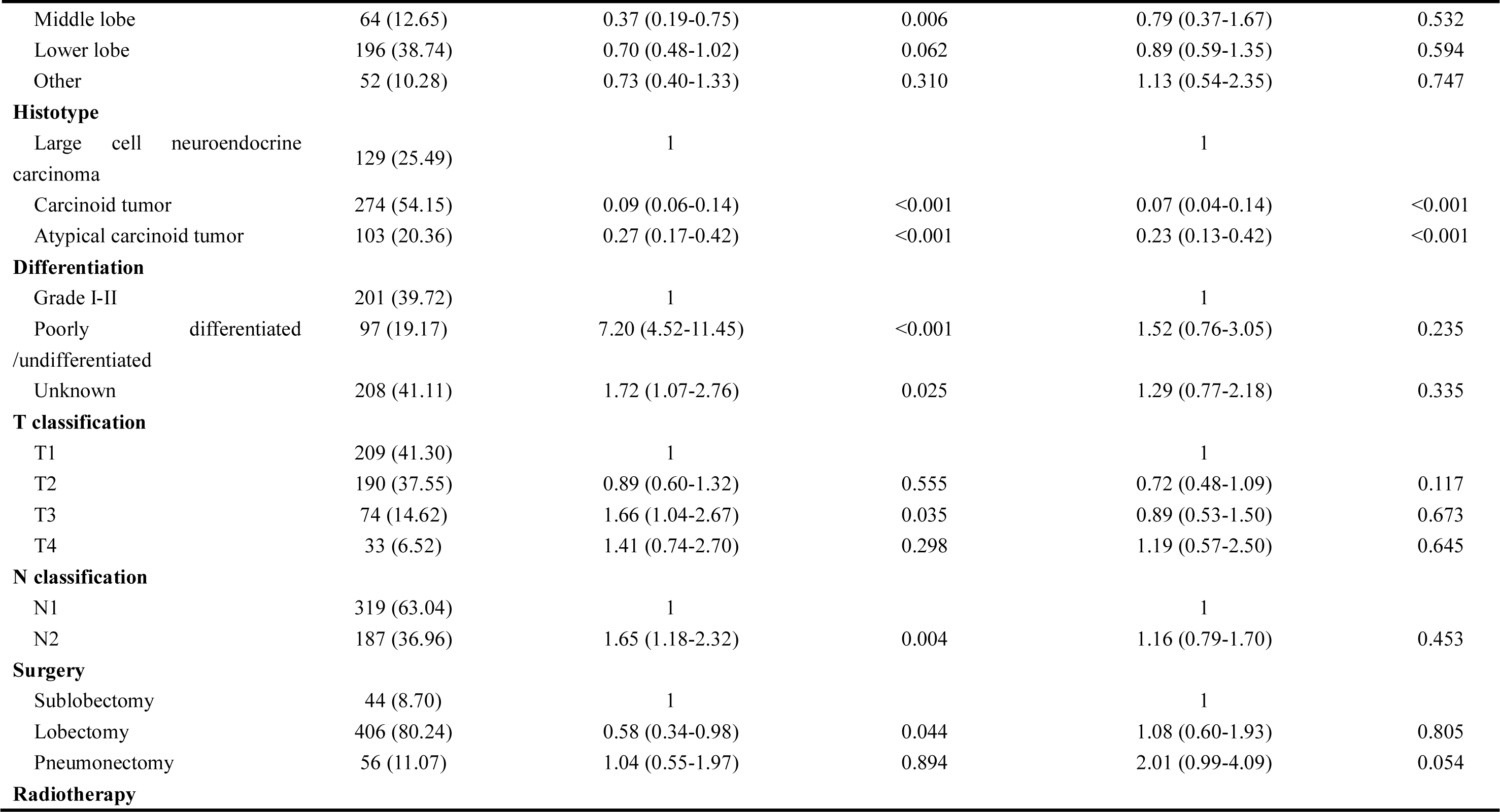
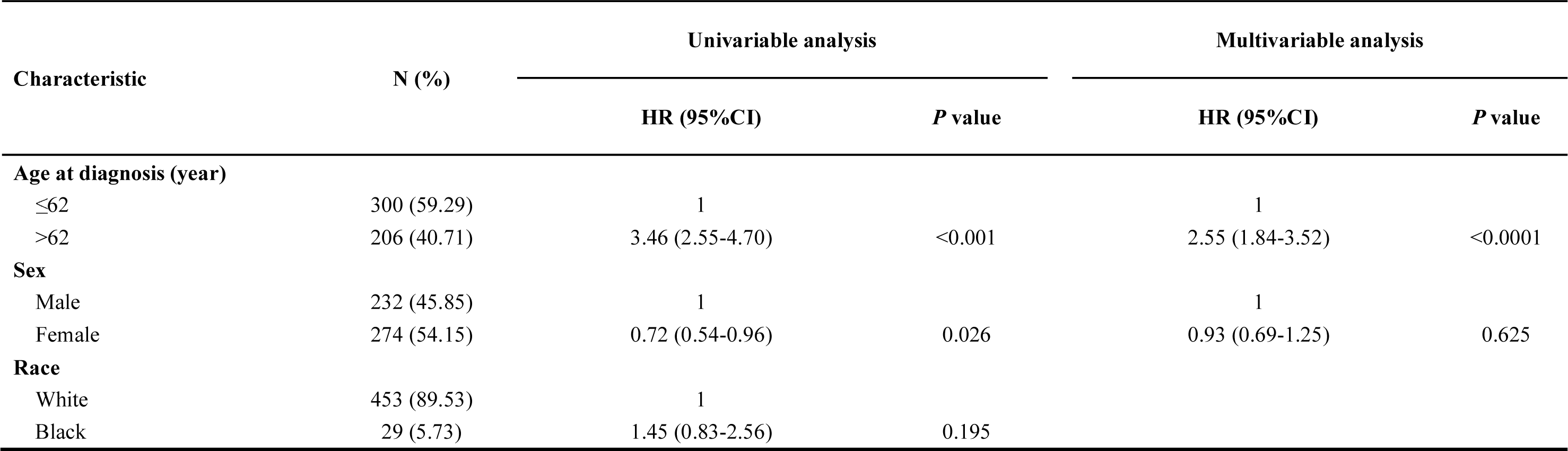
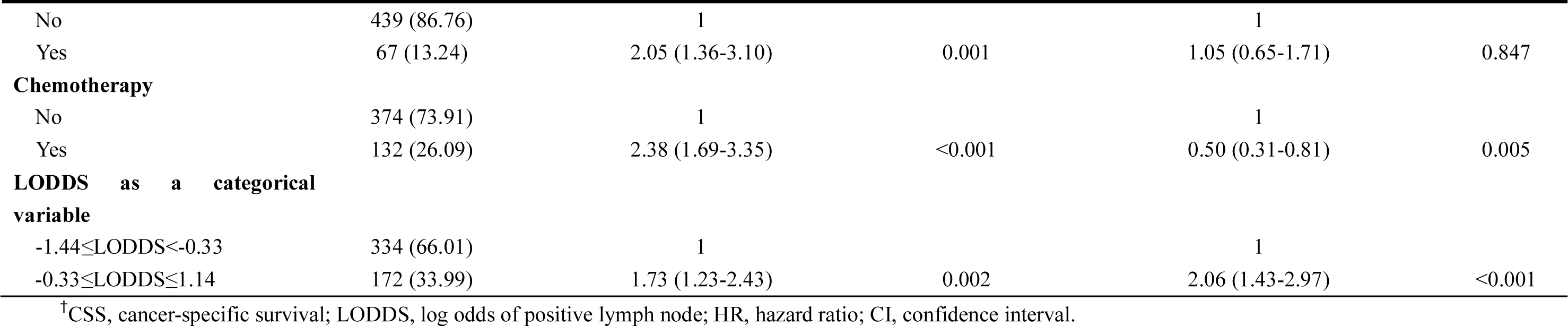
Univariable and multivariable Cox proportional regression analysis for the influence of LODDS o**n CSS of the entire dataset**

**Table S4.**
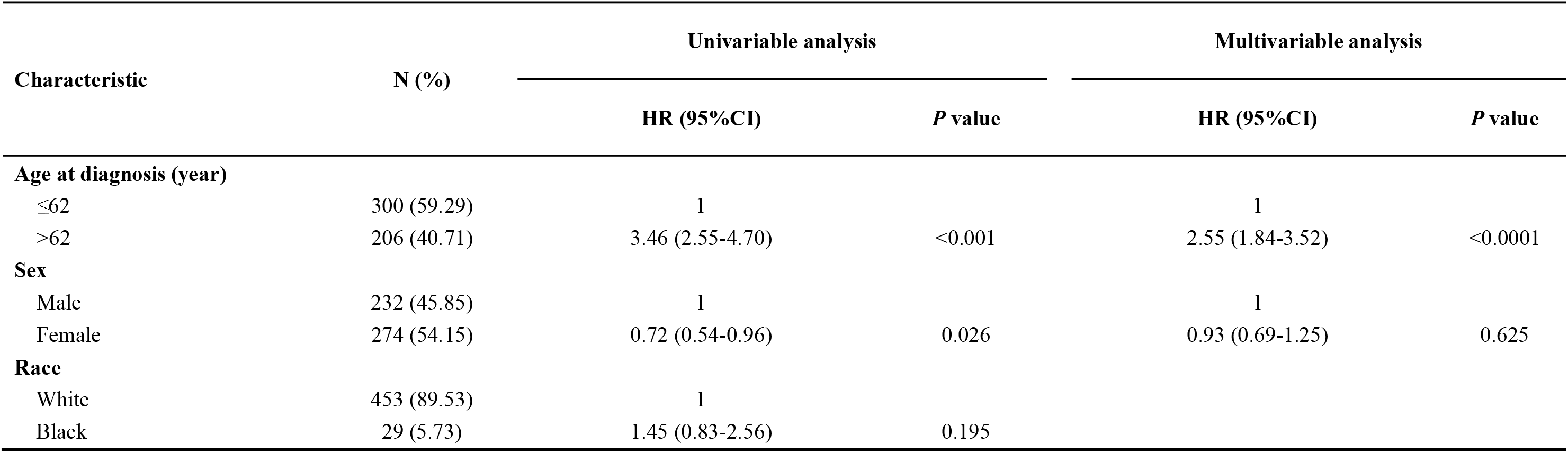
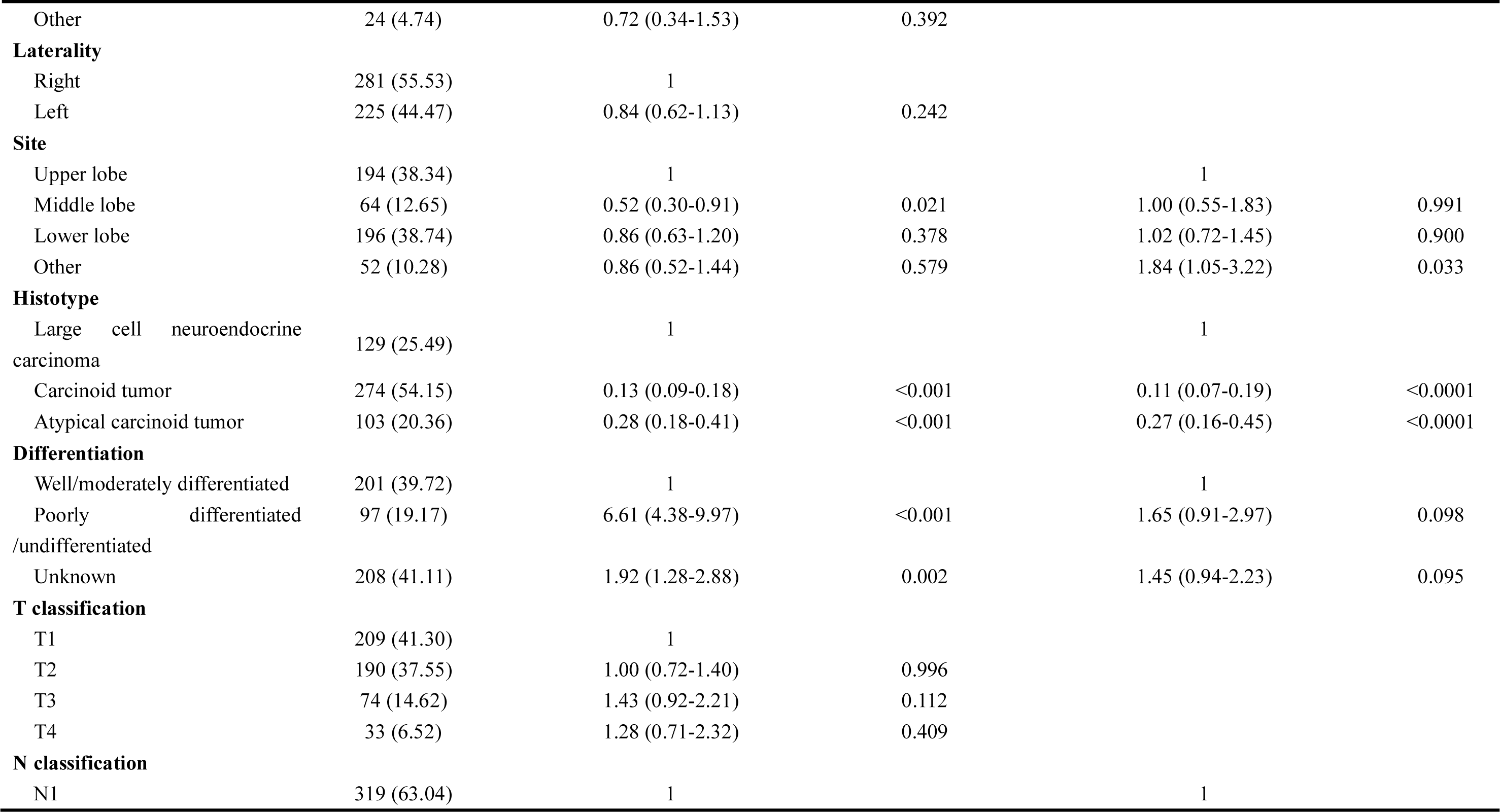
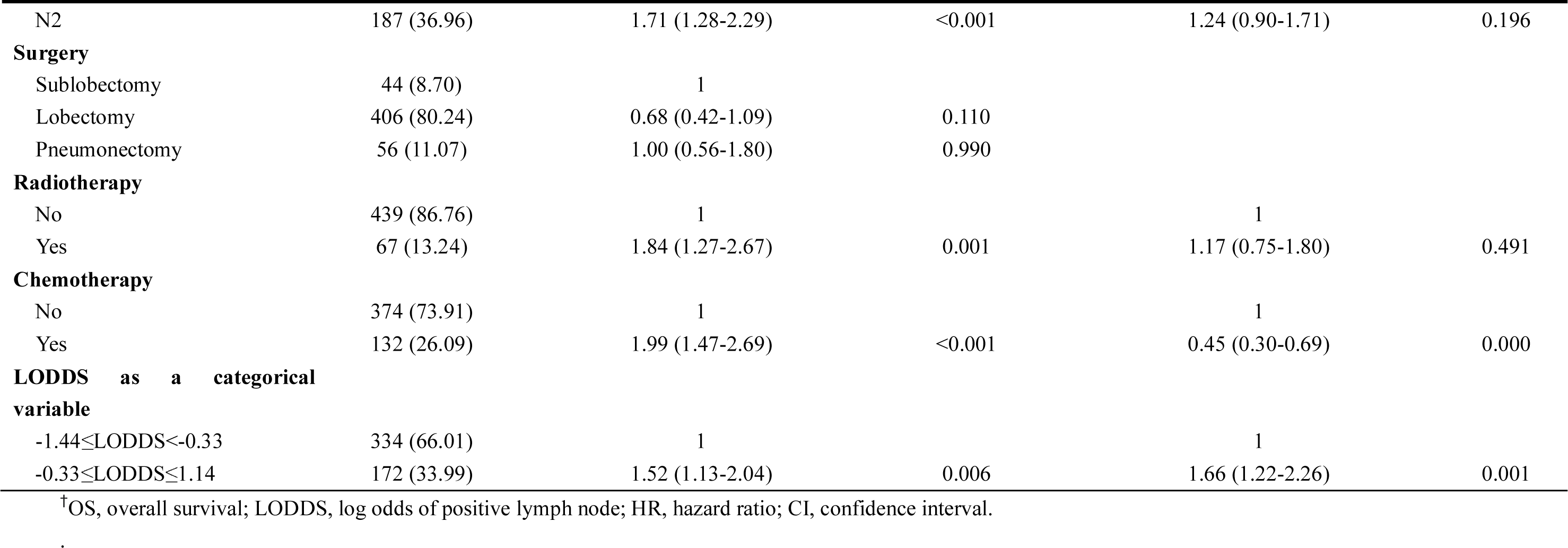
Univariable and multivariable Cox proportional regression analysis for the influence of LODDS on OS of the entire dataset

**Table S5.** Univariable and multivariable Cox proportional regression analysis for the influence of LODDS on CSS of the patients with number of dissected lymph nodes≤20

| Characteristic | N (%) | Univariable analysis |  | Multivariable analysis |  |
| --- | --- | --- | --- | --- | --- |
|  |  | HR (95%CI) | P value | HR (95%CI) | P value |
| **Age at diagnosis (year)** |  |  |  |  |  |
| ≤62 | 269 (59.65) | 1 |  | 1 |  |
| >62 | 182 (40.35) | 2.77 (1.91-4.01) | <0.001 | 2.20 (1.47-3.31) | <0.001 |
| **Sex** |  |  |  |  |  |
| Male | 203 (45.01) | 1 |  |  |  |
| Female | 248 (54.99) | 0.78 (0.55-1.13) | 0.190 |  |  |
| **Race** |  |  |  |  |  |
| White | 405 (89.80) | 1 |  | 1 |  |
| Black | 25 (5.54) | 1.86 (0.97-3.56) | 0.062 | 2.22 (1.09-4.52) | 0.027 |
| Other | 21 (4.66) | 1.02 (0.45-2.32) | 0.970 | 0.67 (0.28-1.59) | 0.367 |
| **Laterality** |  |  |  |  |  |
| Right | 246 (54.55) | 1 |  |  |  |
| Left | 205 (45.45) | 0.80 (0.56-1.16) | 0.242 |  |  |
| **Site** |  |  |  |  |  |
| Upper lobe | 179 (39.69) | 1 |  | 1 |  |
| Middle lobe | 59 (13.08) | 0.32 (0.15-0.70) | 0.004 | 0.66 (0.28-1.57) | 0.351 |
| Lower lobe | 179 (39.69) | 0.67 (0.45-0.99) | 0.043 | 0.97 (0.62-1.53) | 0.912 |
| Other | 34 (7.54) | 0.66 (0.31-1.37) | 0.264 | 1.41 (0.60-3.31) | 0.433 |
| **Histotype** |  |  |  |  |  |
| Large cell neuroendocrine carcinoma | 113 (25.06) | 1 |  | 1 |  |
| Carcinoid tumor | 255 (56.54) | 0.08 (0.05-0.13) | <0.001 | 0.07 (0.03-0.14) | <0.001 |
| Atypical carcinoid tumor | 83 (18.40) | 0.21 (0.13-0.36) | <0.001 | 0.18 (0.09-0.37) | <0.001 |
| **Differentiation** |  |  |  |  |  |
| Well/moderately differentiated | 178 (39.47) | 1 |  | 1 |  |
| Poorly differentiated/undifferentiated | 84 (18.63) | 9.54 (5.65-16.13) | <0.001 | 1.76 (0.81-3.81) | 0.154 |
| Unknown | 189 (41.91) | 2.12 (1.24-3.64) | 0.006 | 1.50 (0.82-2.73) | 0.187 |
| **T classification** |  |  |  |  |  |
| T1 | 193 (42.79) | 1 |  | 1 |  |
| T2 | 170 (37.69) | 0.91 (0.60-1.39) | 0.660 | 0.67 (0.43-1.04) | 0.073 |
| T3 | 62 (13.75) | 1.63 (0.97-2.75) | 0.066 | 0.78 (0.44-1.38) | 0.387 |
| T4 | 26 (5.76) | 1.51 (0.74-3.08) | 0.256 | 1.40 (0.64-3.08) | 0.402 |
| **N classification** |  |  |  |  |  |
| N1 | 283 (62.75) | 1 |  | 1 |  |
| N2 | 168 (37.25) | 1.56 (1.09-2.25) | 0.016 | 1.07 (0.71-1.62) | 0.729 |
| **Surgery** |  |  |  |  |  |
| Sublobectomy | 43 (9.53) | 1 |  | 1 |  |
| Lobectomy | 365 (80.93) | 0.62 (0.36-1.07) | 0.085 | 1.17 (0.64-2.13) | 0.620 |
| Pneumonectomy | 43 (9.53) | 0.84 (0.41-1.71) | 0.626 | 1.66 (0.75-3.67) | 0.207 |
| **Radiotherapy** |  |  |  |  |  |
| No | 387 (85.81) | 1 |  | 1 |  |
| Yes | 64 (14.19) | 2.24 (1.47-3.41) | <0.001 | 1.18 (0.71-1.94) | 0.520 |
| **Chemotherapy** |  |  |  |  |  |
| No | 336 (74.50) | 1 |  | 1 |  |
| Yes | 115 (25.50) | 2.60 (1.80-3.75) | <0.001 | 0.46 (0.27-0.77) | 0.004 |
| **LODDS as a categorical variable** |  |  |  |  |  |
| -1.44≤LODDS<-0.33 | 286 (63.41) | 1 |  | 1 |  |
| -0.33≤LODDS≤1.14 | 165 (36.59) | 1.89 (1.32-2.72) | 0.001 | 2.16 (1.46-3.19) | <0.001 |

<sup>†</sup>CSS, cancer-specific survival; LODDS, log odds of positive lymph node; HR, hazard ratio; CI, confidence interval.

**Table S6.**
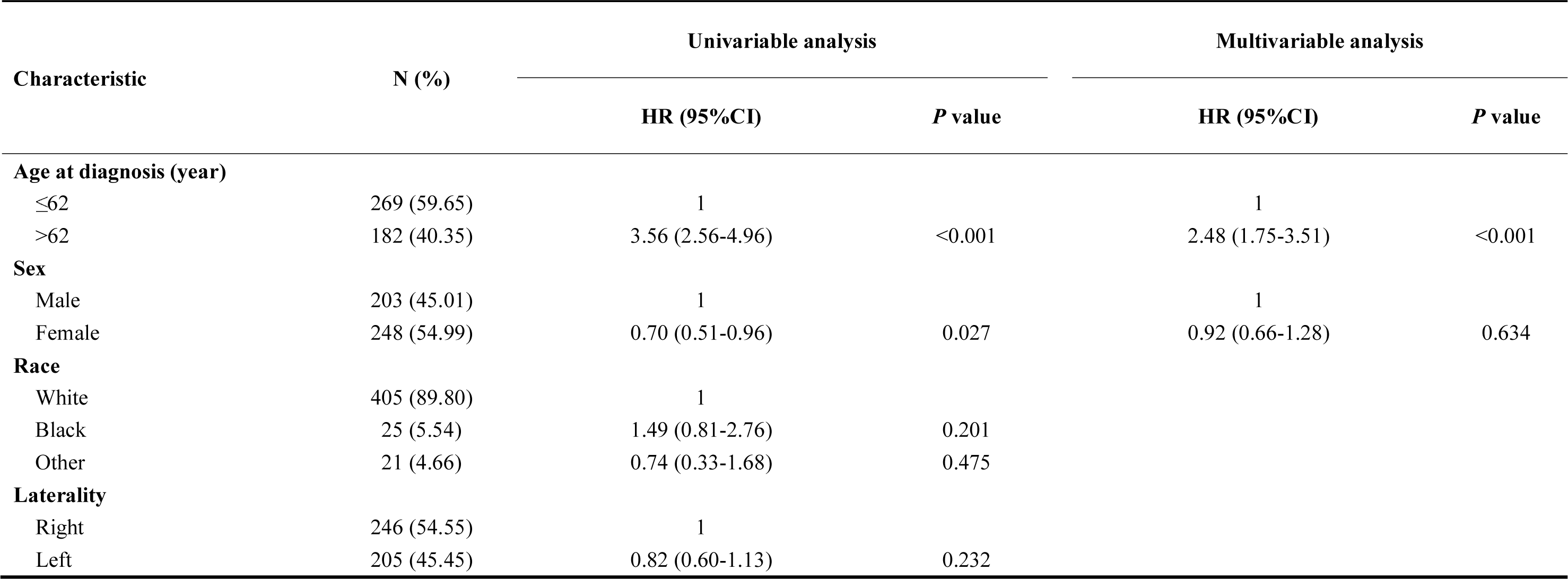
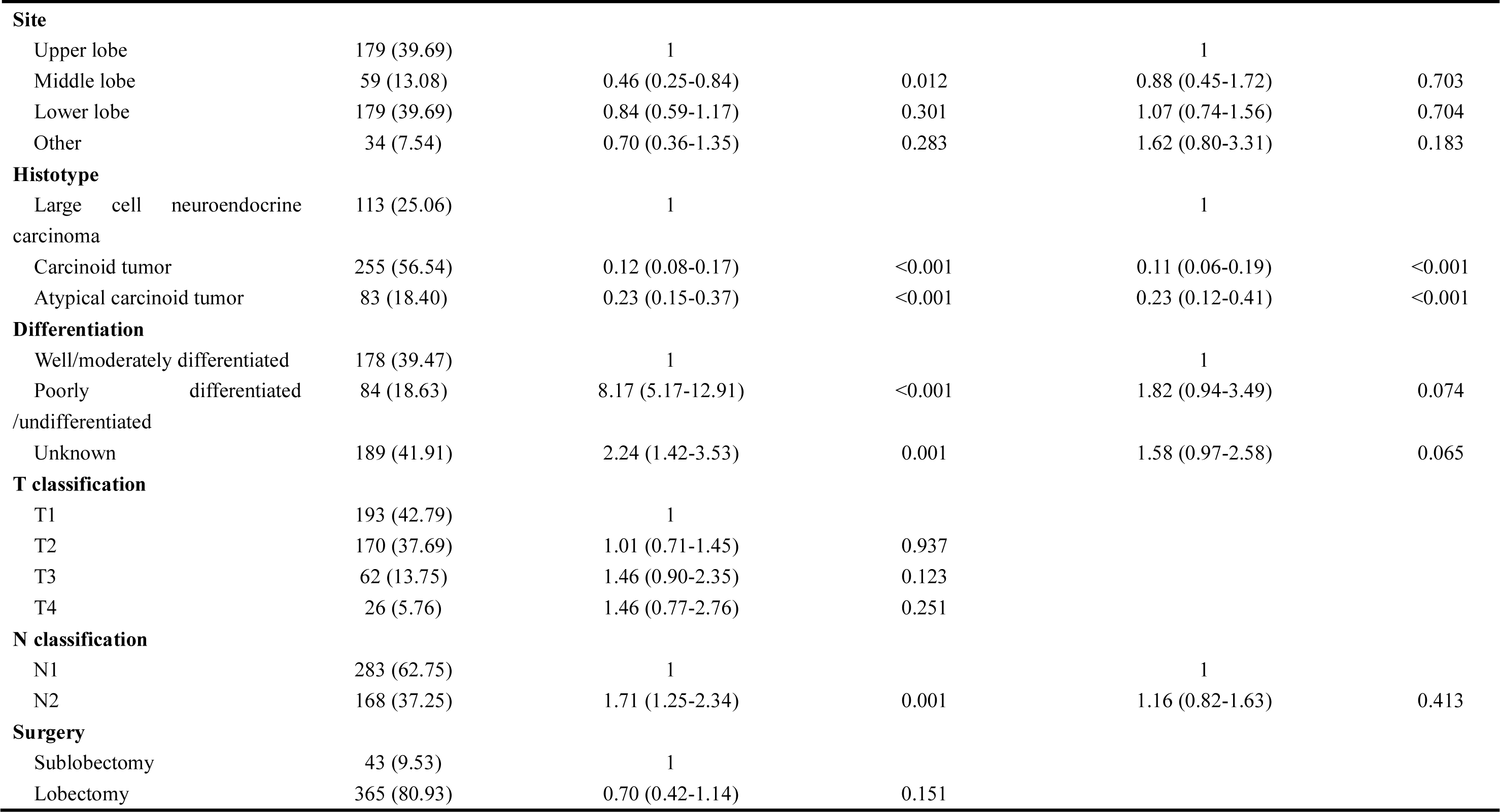
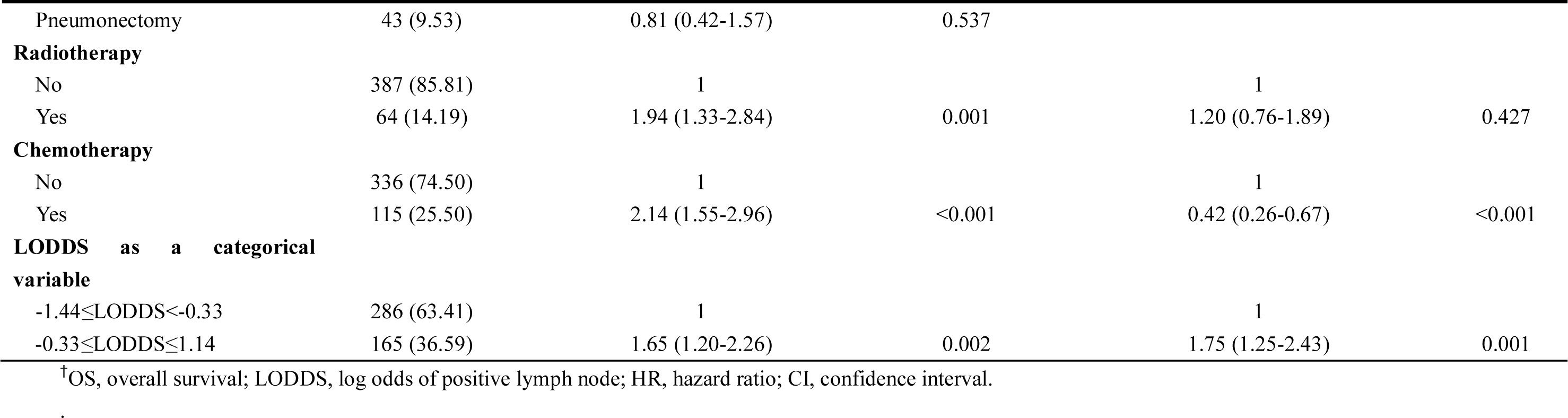
Univariable and multivariable Cox proportional regression analysis for the influence of LODDS on OS of the patients with number of dissected lymph nodes≤20

**Table S7.**
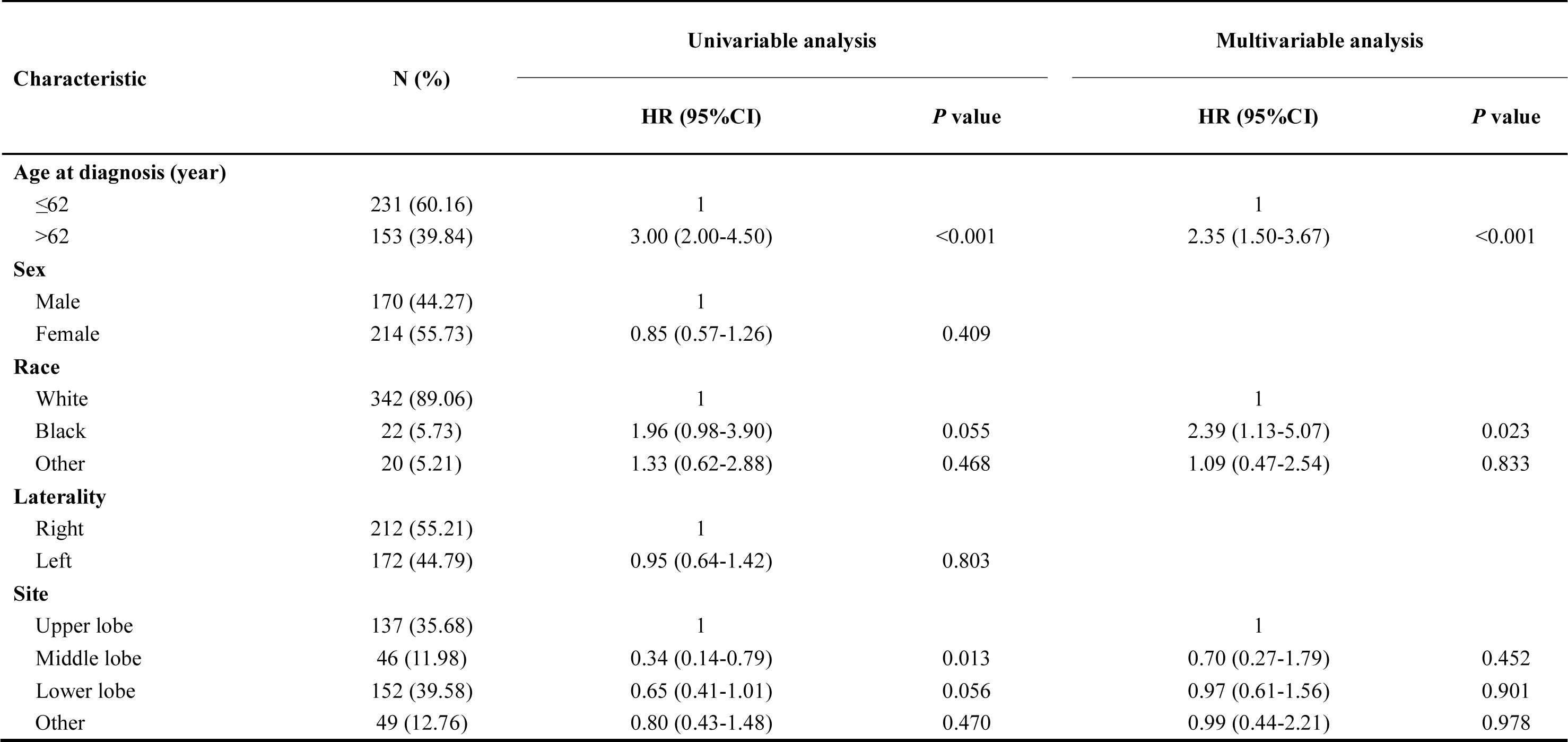
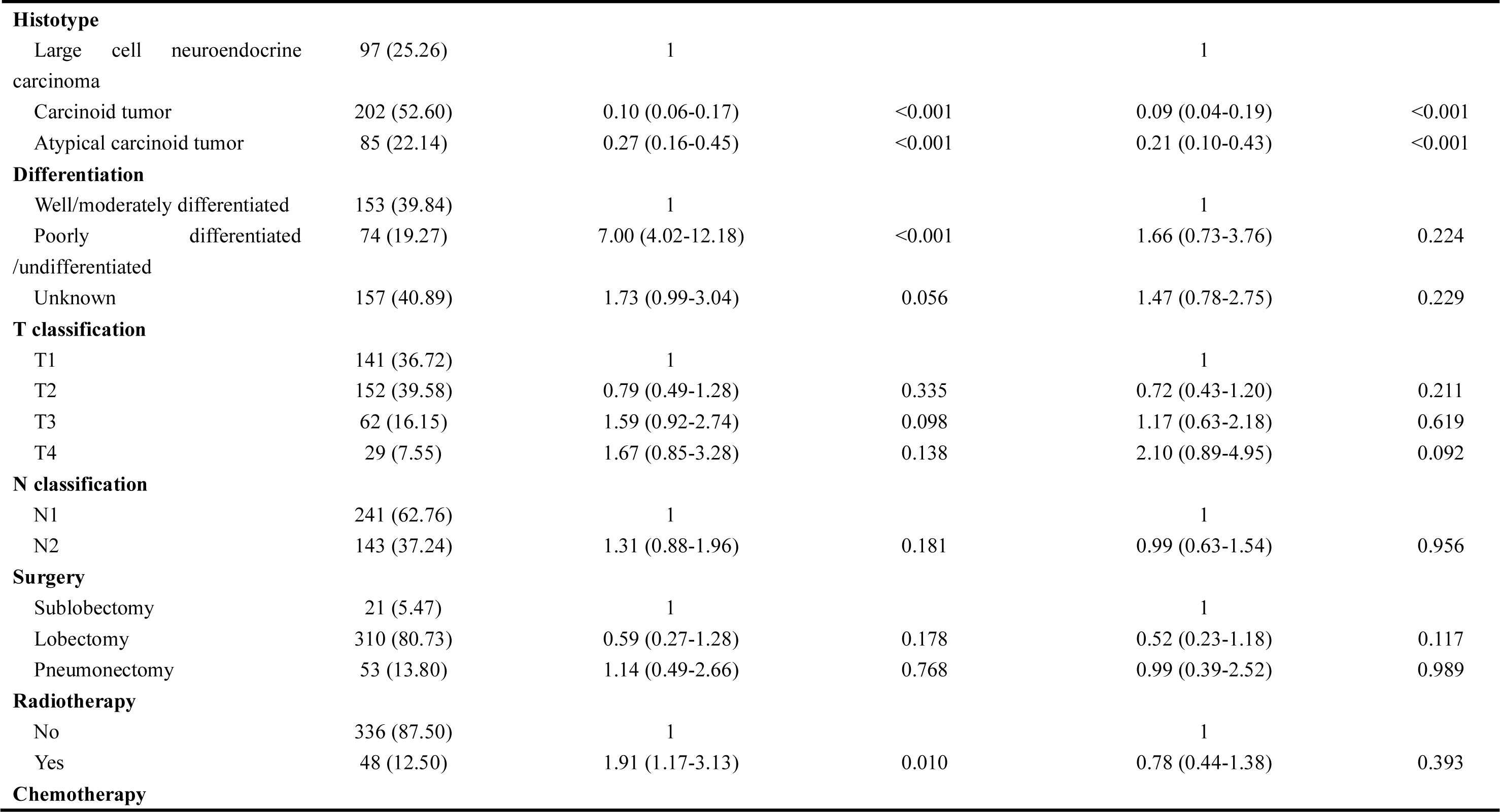
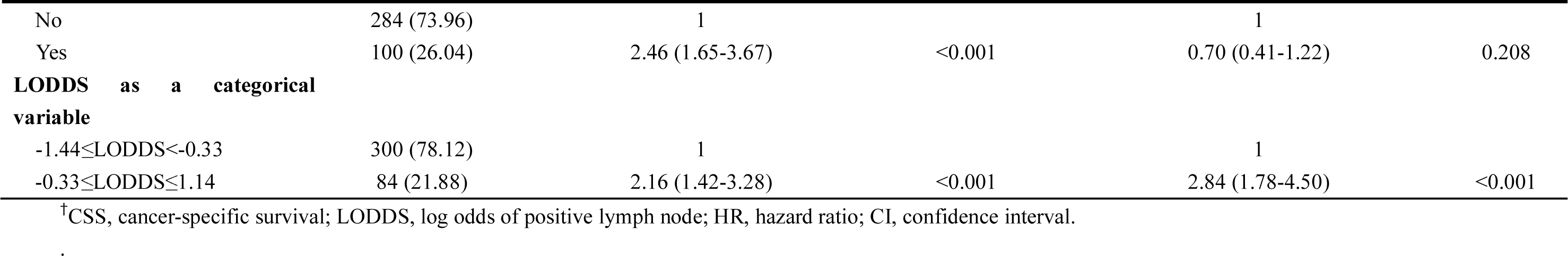
Univariable and multivariable Cox proportional regression analysis for the influence of LODDS on CSS of the patients with number of dissected lymph nodes≥6

| Characteristic | N (%) | Univariable analysis |  | Multivariable analysis |  |
| --- | --- | --- | --- | --- | --- |
|  |  | HR (95%CI) | P value | HR (95%CI) | P value |
| Age at diagnosis (year) |  |  |  |  |  |
| ≤62 | 231 (60.16) | 1 |  | 1 |  |
| >62 | 153 (39.84) | 3.00 (2.00-4.50) | <0.001 | 2.35 (1.50-3.67) | <0.001 |
| Sex |  |  |  |  |  |
| Male | 170 (44.27) | 1 |  |  |  |
| Female | 214 (55.73) | 0.85 (0.57-1.26) | 0.409 |  |  |
| Race |  |  |  |  |  |
| White | 342 (89.06) | 1 |  | 1 |  |
| Black | 22 (5.73) | 1.96 (0.98-3.90) | 0.055 | 2.39 (1.13-5.07) | 0.023 |
| Other | 20 (5.21) | 1.33 (0.62-2.88) | 0.468 | 1.09 (0.47-2.54) | 0.833 |
| Laterality |  |  |  |  |  |
| Right | 212 (55.21) | 1 |  |  |  |
| Left | 172 (44.79) | 0.95 (0.64-1.42) | 0.803 |  |  |
| Site |  |  |  |  |  |
| Upper lobe | 137 (35.68) | 1 |  | 1 |  |
| Middle lobe | 46 (11.98) | 0.34 (0.14-0.79) | 0.013 | 0.70 (0.27-1.79) | 0.452 |
| Lower lobe | 152 (39.58) | 0.65 (0.41-1.01) | 0.056 | 0.97 (0.61-1.56) | 0.901 |
| Other | 49 (12.76) | 0.80 (0.43-1.48) | 0.470 | 0.99 (0.44-2.21) | 0.978 |
| **Histotype** |  |  |  |  |  |
| Large cell neuroendocrine carcinoma | 97 (25.26) | 1 |  | 1 |  |
| Carcinoid tumor | 202 (52.60) | 0.10 (0.06-0.17) | <0.001 | 0.09 (0.04-0.19) | <0.001 |
| Atypical carcinoid tumor | 85 (22.14) | 0.27 (0.16-0.45) | <0.001 | 0.21 (0.10-0.43) | <0.001 |
| **Differentiation** |  |  |  |  |  |
| Well/moderately differentiated | 153 (39.84) | 1 |  | 1 |  |
| Poorly differentiated | 74 (19.27) | 7.00 (4.02-12.18) | <0.001 | 1.66 (0.73-3.76) | 0.224 |
| /undifferentiated |  |  |  |  |  |
| Unknown | 157 (40.89) | 1.73 (0.99-3.04) | 0.056 | 1.47 (0.78-2.75) | 0.229 |
| **T classification** |  |  |  |  |  |
| T1 | 141 (36.72) | 1 |  | 1 |  |
| T2 | 152 (39.58) | 0.79 (0.49-1.28) | 0.335 | 0.72 (0.43-1.20) | 0.211 |
| T3 | 62 (16.15) | 1.59 (0.92-2.74) | 0.098 | 1.17 (0.63-2.18) | 0.619 |
| T4 | 29 (7.55) | 1.67 (0.85-3.28) | 0.138 | 2.10 (0.89-4.95) | 0.092 |
| **N classification** |  |  |  |  |  |
| N1 | 241 (62.76) | 1 |  | 1 |  |
| N2 | 143 (37.24) | 1.31 (0.88-1.96) | 0.181 | 0.99 (0.63-1.54) | 0.956 |
| **Surgery** |  |  |  |  |  |
| Sublobectomy | 21 (5.47) | 1 |  | 1 |  |
| Lobectomy | 310 (80.73) | 0.59 (0.27-1.28) | 0.178 | 0.52 (0.23-1.18) | 0.117 |
| Pneumonectomy | 53 (13.80) | 1.14 (0.49-2.66) | 0.768 | 0.99 (0.39-2.52) | 0.989 |
| **Radiotherapy** |  |  |  |  |  |
| No | 336 (87.50) | 1 |  | 1 |  |
| Yes | 48 (12.50) | 1.91 (1.17-3.13) | 0.010 | 0.78 (0.44-1.38) | 0.393 |
| **Chemotherapy** |  |  |  |  |  |
| No | 284 (73.96) | 1 |  | 1 |  |
| Yes | 100 (26.04) | 2.46 (1.65-3.67) | <0.001 | 0.70 (0.41-1.22) | 0.208 |
| **LODDS as a categorical variable** |  |  |  |  |  |
| -1.44≤LODDS<-0.33 | 300 (78.12) | 1 |  | 1 |  |
| -0.33≤LODDS≤1.14 | 84 (21.88) | 2.16 (1.42-3.28) | <0.001 | 2.84 (1.78-4.50) | <0.001 |

<sup>†</sup>CSS, cancer-specific survival; LODDS, log odds of positive lymph node; HR, hazard ratio; CI, confidence interval.

**Table S8.**
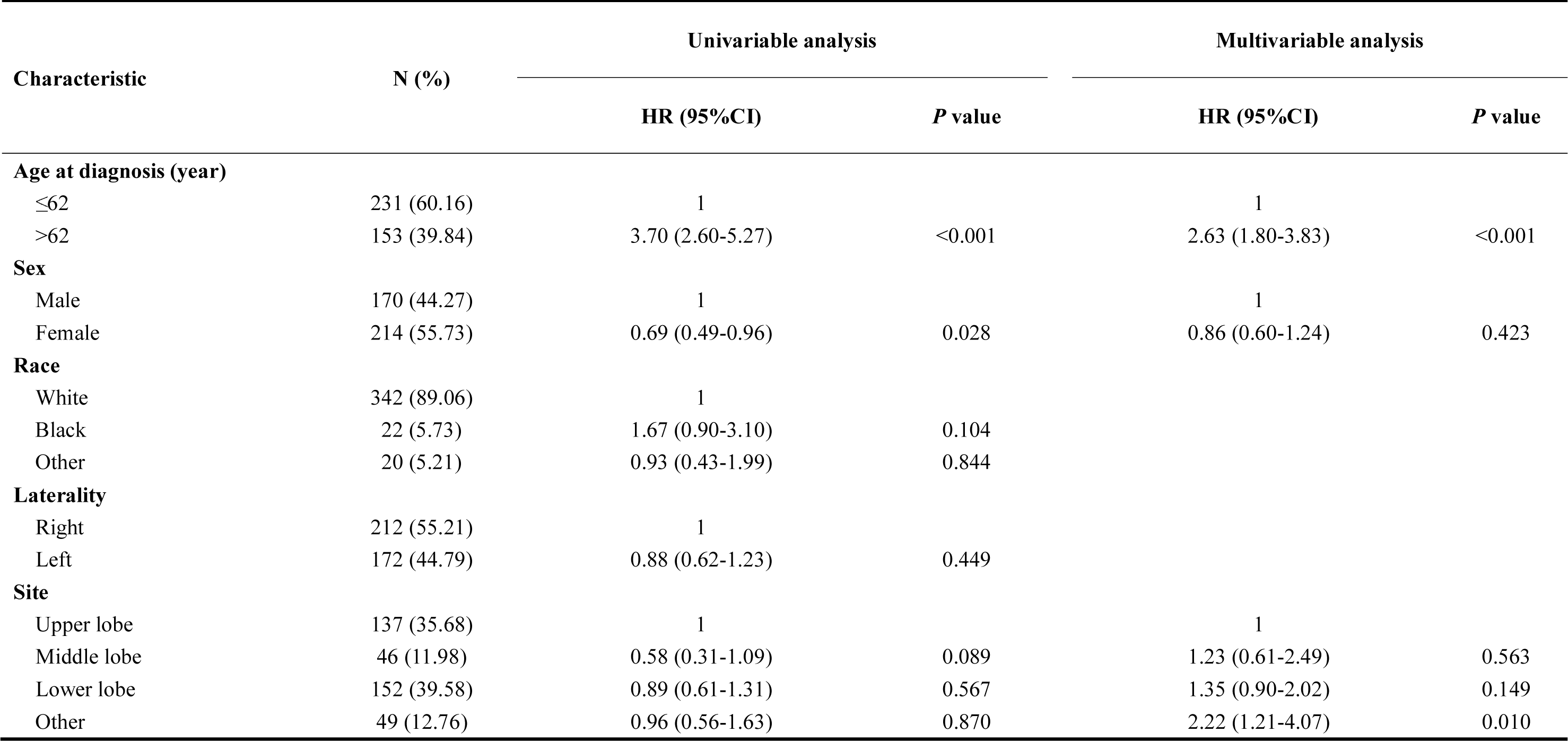
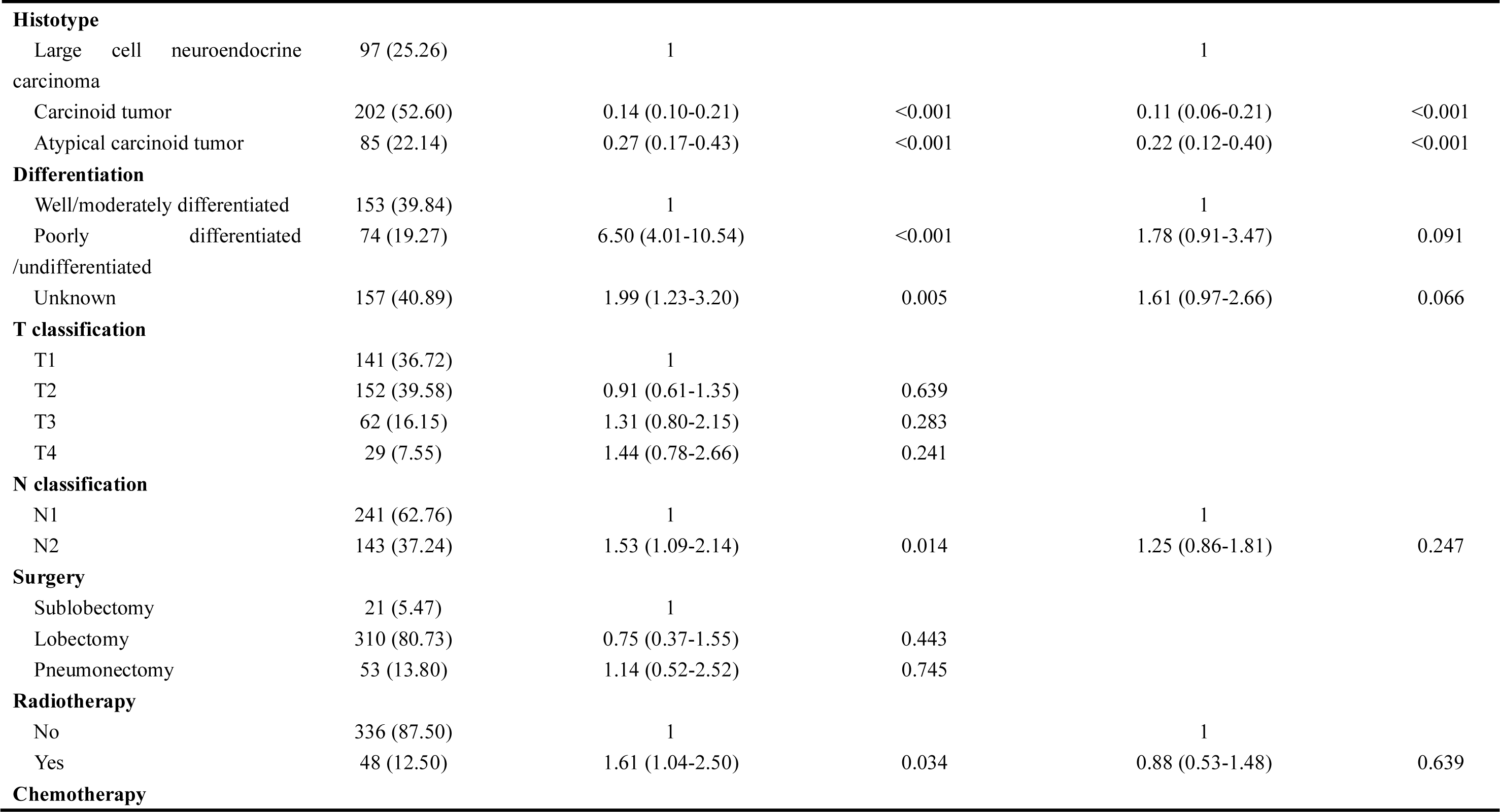
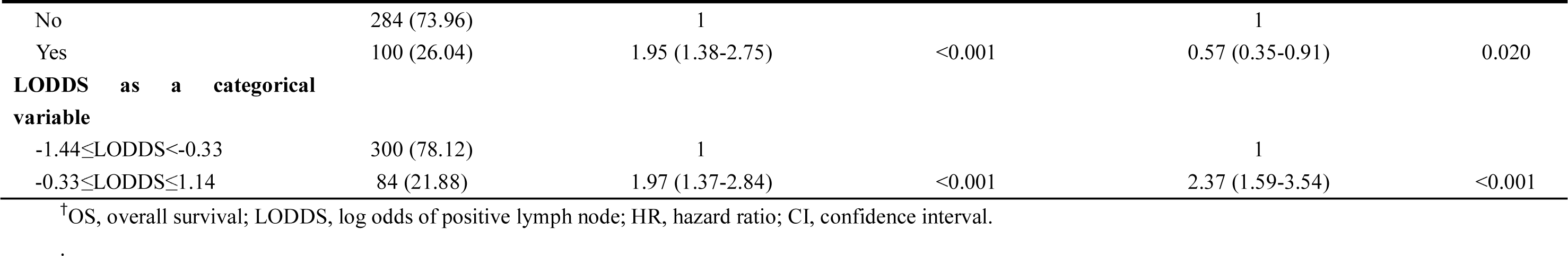
Univariable and multivariable Cox proportional regression analysis for the influence of LODDS on OS of the patients with number of dissected lymph nodes≥6

| Characteristic | N (%) | Univariable analysis |  | Multivariable analysis |  |
| --- | --- | --- | --- | --- | --- |
|  |  | HR (95%CI) | P value | HR (95%CI) | P value |
| Age at diagnosis (year) |  |  |  |  |  |
| ≤62 | 231 (60.16) | 1 |  | 1 |  |
| >62 | 153 (39.84) | 3.70 (2.60-5.27) | <0.001 | 2.63 (1.80-3.83) | <0.001 |
| Sex |  |  |  |  |  |
| Male | 170 (44.27) | 1 |  | 1 |  |
| Female | 214 (55.73) | 0.69 (0.49-0.96) | 0.028 | 0.86 (0.60-1.24) | 0.423 |
| Race |  |  |  |  |  |
| White | 342 (89.06) | 1 |  |  |  |
| Black | 22 (5.73) | 1.67 (0.90-3.10) | 0.104 |  |  |
| Other | 20 (5.21) | 0.93 (0.43-1.99) | 0.844 |  |  |
| Laterality |  |  |  |  |  |
| Right | 212 (55.21) | 1 |  |  |  |
| Left | 172 (44.79) | 0.88 (0.62-1.23) | 0.449 |  |  |
| Site |  |  |  |  |  |
| Upper lobe | 137 (35.68) | 1 |  | 1 |  |
| Middle lobe | 46 (11.98) | 0.58 (0.31-1.09) | 0.089 | 1.23 (0.61-2.49) | 0.563 |
| Lower lobe | 152 (39.58) | 0.89 (0.61-1.31) | 0.567 | 1.35 (0.90-2.02) | 0.149 |
| Other | 49 (12.76) | 0.96 (0.56-1.63) | 0.870 | 2.22 (1.21-4.07) | 0.010 |
| **Histotype** |  |  |  |  |  |
| Large cell neuroendocrine carcinoma | 97 (25.26) | 1 |  | 1 |  |
| Carcinoid tumor | 202 (52.60) | 0.14 (0.10-0.21) | <0.001 | 0.11 (0.06-0.21) | <0.001 |
| Atypical carcinoid tumor | 85 (22.14) | 0.27 (0.17-0.43) | <0.001 | 0.22 (0.12-0.40) | <0.001 |
| **Differentiation** |  |  |  |  |  |
| Well/moderately differentiated | 153 (39.84) | 1 |  | 1 |  |
| Poorly differentiated | 74 (19.27) | 6.50 (4.01-10.54) | <0.001 | 1.78 (0.91-3.47) | 0.091 |
| /undifferentiated |  |  |  |  |  |
| Unknown | 157 (40.89) | 1.99 (1.23-3.20) | 0.005 | 1.61 (0.97-2.66) | 0.066 |
| **T classification** |  |  |  |  |  |
| T1 | 141 (36.72) | 1 |  |  |  |
| T2 | 152 (39.58) | 0.91 (0.61-1.35) | 0.639 |  |  |
| T3 | 62 (16.15) | 1.31 (0.80-2.15) | 0.283 |  |  |
| T4 | 29 (7.55) | 1.44 (0.78-2.66) | 0.241 |  |  |
| **N classification** |  |  |  |  |  |
| N1 | 241 (62.76) | 1 |  | 1 |  |
| N2 | 143 (37.24) | 1.53 (1.09-2.14) | 0.014 | 1.25 (0.86-1.81) | 0.247 |
| **Surgery** |  |  |  |  |  |
| Sublobectomy | 21 (5.47) | 1 |  |  |  |
| Lobectomy | 310 (80.73) | 0.75 (0.37-1.55) | 0.443 |  |  |
| Pneumonectomy | 53 (13.80) | 1.14 (0.52-2.52) | 0.745 |  |  |
| **Radiotherapy** |  |  |  |  |  |
| No | 336 (87.50) | 1 |  | 1 |  |
| Yes | 48 (12.50) | 1.61 (1.04-2.50) | 0.034 | 0.88 (0.53-1.48) | 0.639 |
| **Chemotherapy** |  |  |  |  |  |
| No | 284 (73.96) | 1 |  | 1 |  |
| Yes | 100 (26.04) | 1.95 (1.38-2.75) | <0.001 | 0.57 (0.35-0.91) | 0.020 |
| **LODDS as a categorical variable** |  |  |  |  |  |
| -1.44≤LODDS<-0.33 | 300 (78.12) | 1 |  | 1 |  |
| -0.33≤LODDS≤1.14 | 84 (21.88) | 1.97 (1.37-2.84) | <0.001 | 2.37 (1.59-3.54) | <0.001 |

<sup>†</sup>OS, overall survival; LODDS, log odds of positive lymph node; HR, hazard ratio; CI, confidence interval.

**Table S9.** HRs for subgroups

| Subgroups | N (%) | CSS |  | OS |  |
| --- | --- | --- | --- | --- | --- |
|  |  | HR (95%CI) | P value | HR (95%CI) | P value |
| Age at diagnosis (year) |  |  |  |  |  |
| ≤62 | 300 (59.29) | 2.52 (1.42-4.47) | 0.002 | 2.20 (1.34-3.63) | 0.002 |
| >62 | 206 (40.71) | 1.63 (0.98-2.69) | 0.058 | 1.21 (0.80-1.82) | 0.369 |
| Sex |  |  |  |  |  |
| Male | 232 (45.85) | 3.96 (2.15-7.32) | <0.001 | 2.06 (1.29-3.27) | 0.002 |
| Female | 274 (54.15) | 1.53 (0.90-2.58) | 0.116 | 1.43 (0.92-2.23) | 0.110 |
| Laterality |  |  |  |  |  |
| Right | 281 (55.53) | 1.67 (1.04-2.67) | 0.035 | 1.56 (1.04-2.35) | 0.033 |
| Left | 225 (44.47) | 2.88 (1.52-5.45) | 0.001 | 2.00 (1.20-3.32) | 0.008 |
| Site |  |  |  |  |  |
| Pulmonary lobe | 454 (89.72) | 2.10 (1.44-3.08) | <0.001 | 1.67 (1.21-2.31) | 0.002 |
| Other | 52 (10.28) | 4.65 (0.25-87.17) | 0.304 | 1.45 (0.37-5.73) | 0.594 |
| Histotype |  |  |  |  |  |
| Large cell neuroendocrine carcinoma | 129 (25.49) | 1.96 (1.19-3.24) | 0.009 | 1.75 (1.12-2.74) | 0.014 |
| Carcinoid tumor | 274 (54.15) | 0.75 (0.32-1.73) | 0.500 | 0.79 (0.43-1.43) | 0.435 |
| Atypical carcinoid tumor | 103 (20.36) | 2.67 (0.90-7.90) | 0.076 | 2.40 (1.01-5.74) | 0.048 |
| Differentiation |  |  |  |  |  |
| Well/moderately differentiated | 201 (39.72) | 0.70 (0.27-1.80) | 0.455 | 0.79 (0.37-1.70) | 0.551 |
| Poorly differentiated | 97 (19.17) | 1.96 (1.06-3.61) | 0.031 | 1.45 (0.85-2.48) | 0.174 |
| /undifferentiated |  |  |  |  |  |
| Unknown | 208 (41.11) | 3.38 (1.70-6.70) | <0.001 | 2.31 (1.40-3.82) | 0.001 |
| **T classification** |  |  |  |  |  |
| T1 | 209 (41.30) | 2.10 (1.15-3.84) | 0.016 | 1.65 (0.98-2.77) | 0.058 |
| T2 | 190 (37.55) | 1.95 (1.00-3.80) | 0.048 | 1.36 (0.81-2.28) | 0.243 |
| T3-4 | 107 (21.15) | 1.75 (0.72-4.22) | 0.217 | 1.28 (0.59-2.77) | 0.530 |
| **N classification** |  |  |  |  |  |
| N1 | 319 (63.04) | 1.99 (1.14-3.45) | 0.015 | 1.56 (0.98-2.49) | 0.060 |
| N2 | 187 (36.96) | 2.24 (1.27-3.96) | 0.006 | 1.87 (1.20-2.93) | 0.006 |
| **Surgery** |  |  |  |  |  |
| Sublobectomy | 44 (8.70) | 4.64 (0.37-57.53) | 0.233 | 0.57 (0.17-1.92) | 0.366 |
| Lobectomy | 406 (80.24) | 2.53 (1.65-3.89) | <0.001 | 2.03 (1.41-2.92) | <0.001 |
| Pneumonectomy | 56 (11.07) | 2.19 (0.46-10.51) | 0.325 | 1.67 (0.51-5.53) | 0.400 |

<sup>†</sup>HR for CSS were evaluated based on multivariable Cox proportional regression model adjusted by age, race, site, histotype, differentiation, T classification, N classification, surgery, radiotherapy and chemotherapy.
<sup>‡</sup>HR for OS were evaluated based on multivariable Cox proportional regression model adjusted by age, race, site, histotype, differentiation, N classification, radiotherapy and chemotherapy.
<sup>§</sup>CSS, cancer-specific survival; LODDS, log odds of positive lymph node; OS, overall survival; HR, hazard ratio; CI, confidence interval.

**Table S10.**
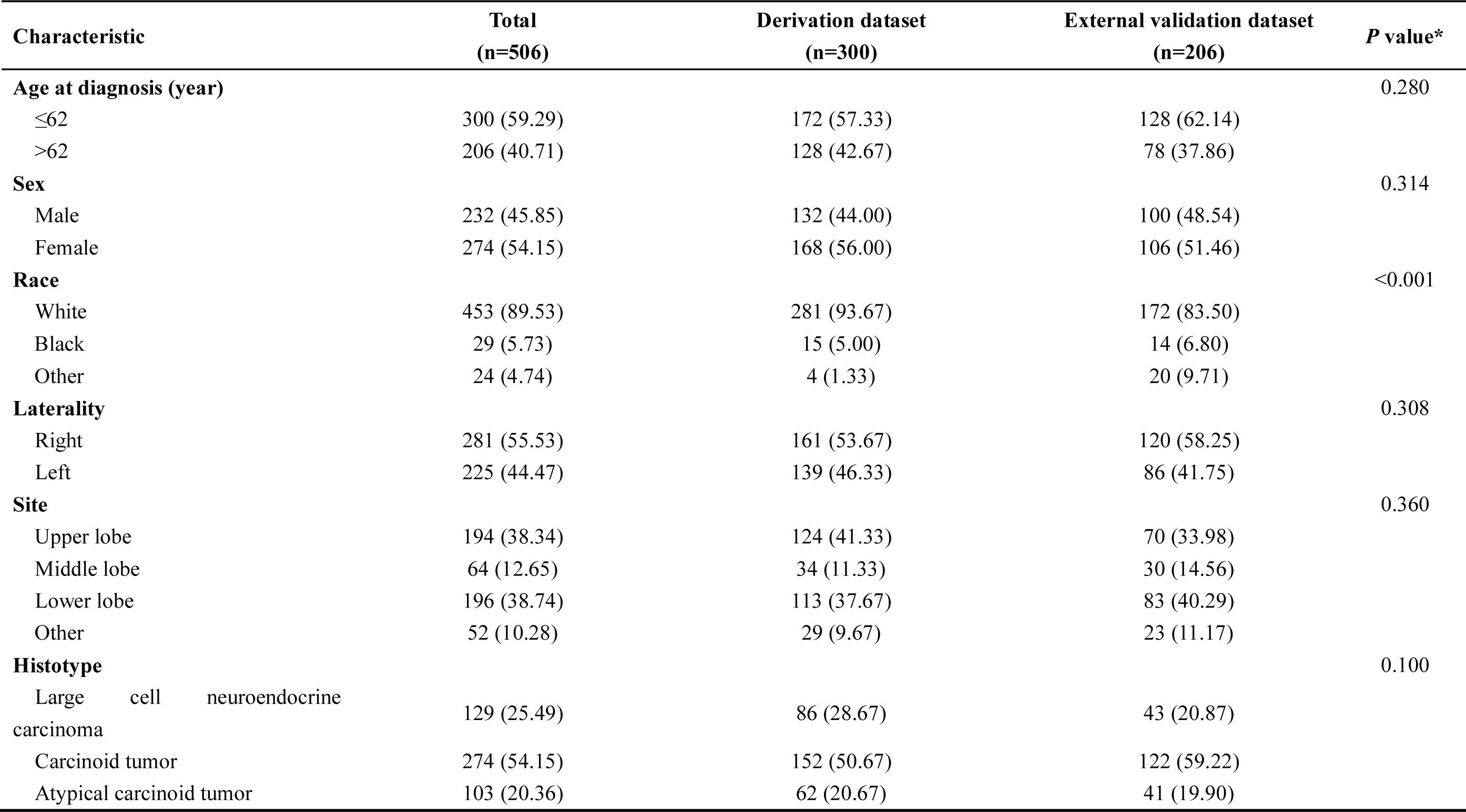
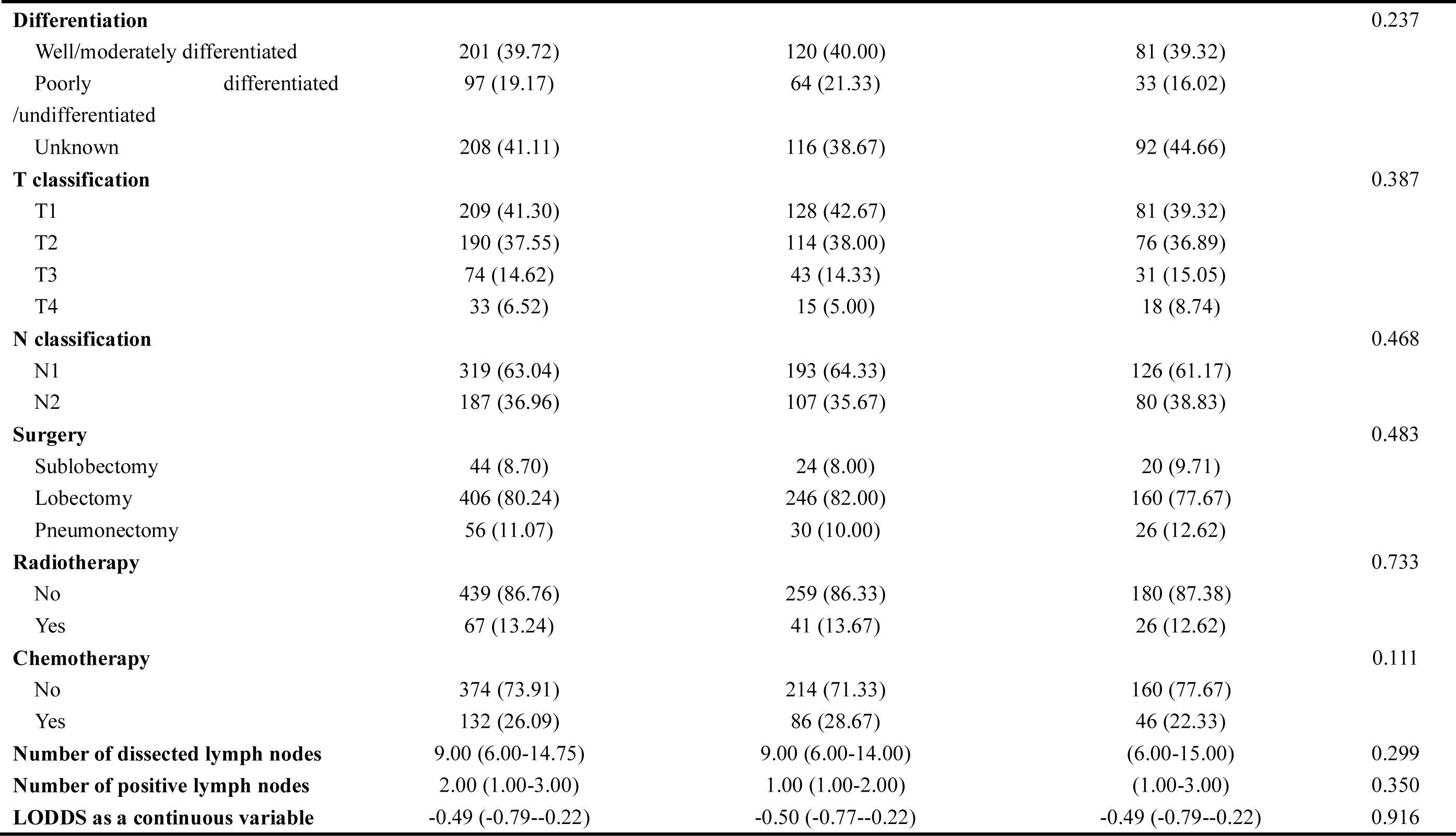
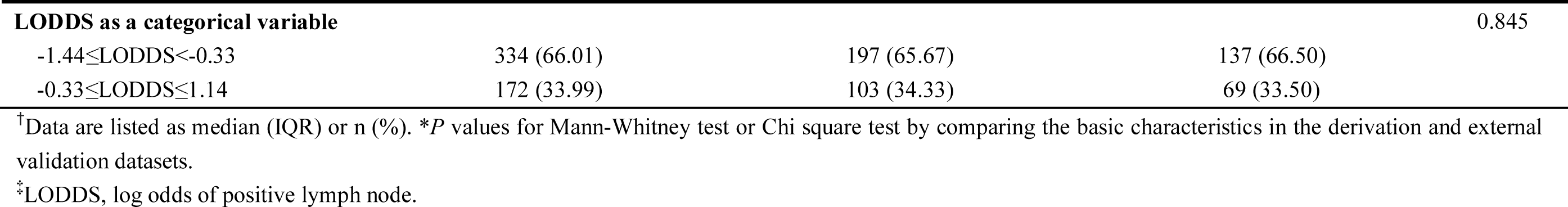
Baseline characteristics of derivation dataset and external validation dataset

**Table S11.** The IDI and continuous-NRI comparing nomogram with AJCC 8th TNM staging system

| Endpoint | IDI |  | Continuous-NRI |  |
| --- | --- | --- | --- | --- |
|  | 95%CI | *P* value | 95%CI | *P* value |
| **Derivation dataset** |  |  |  |  |
| 1-year CSS | 0.249 (0.389-0.161) | <0.001 | 0.721 (0.825-0.554) | <0.001 |
| 3-year CSS | 0.391 (0.519-0.243) | <0.001 | 0.645 (0.743-0.414) | <0.001 |
| 5-year CSS | 0.42 (0.537-0.265) | <0.001 | 0.627 (0.768-0.403) | <0.001 |
| 1-year OS | 0.256 (0.368-0.166) | <0.001 | 0.649 (0.778-0.453) | <0.001 |
| 3-year OS | 0.349 (0.46-0.208) | <0.001 | 0.492 (0.653-0.32) | <0.001 |
| 5-year OS | 0.364 (0.457-0.237) | <0.001 | 0.523 (0.662-0.324) | <0.001 |
| **External validation dataset** |  |  |  |  |
| 1-year CSS | 0.165 (0.356-0.065) | <0.001 | 0.46 (0.683-0.178) | <0.001 |
| 3-year CSS | 0.303 (0.427-0.171) | <0.001 | 0.539 (0.701-0.308) | <0.001 |
| 5-year CSS | 0.321 (0.442-0.146) | <0.001 | 0.496 (0.688-0.283) | <0.001 |
| 1-year OS | 0.121 (0.263-0.057) | <0.001 | 0.379 (0.647-0.162) | <0.001 |
| 3-year OS | 0.257 (0.384-0.138) | <0.001 | 0.456 (0.653-0.23) | <0.001 |
| 5-year OS | 0.274 (0.382-0.11) | <0.001 | 0.451 (0.611-0.24) | <0.001 |

<sup>†</sup>CSS, cancer-specific survival; IDI, integrated discrimination improvement; LODDS, log odds of positive lymph node; NRI, net reclassification improvement; OS, overall survival.

## Figure legends

**Figure S1.**
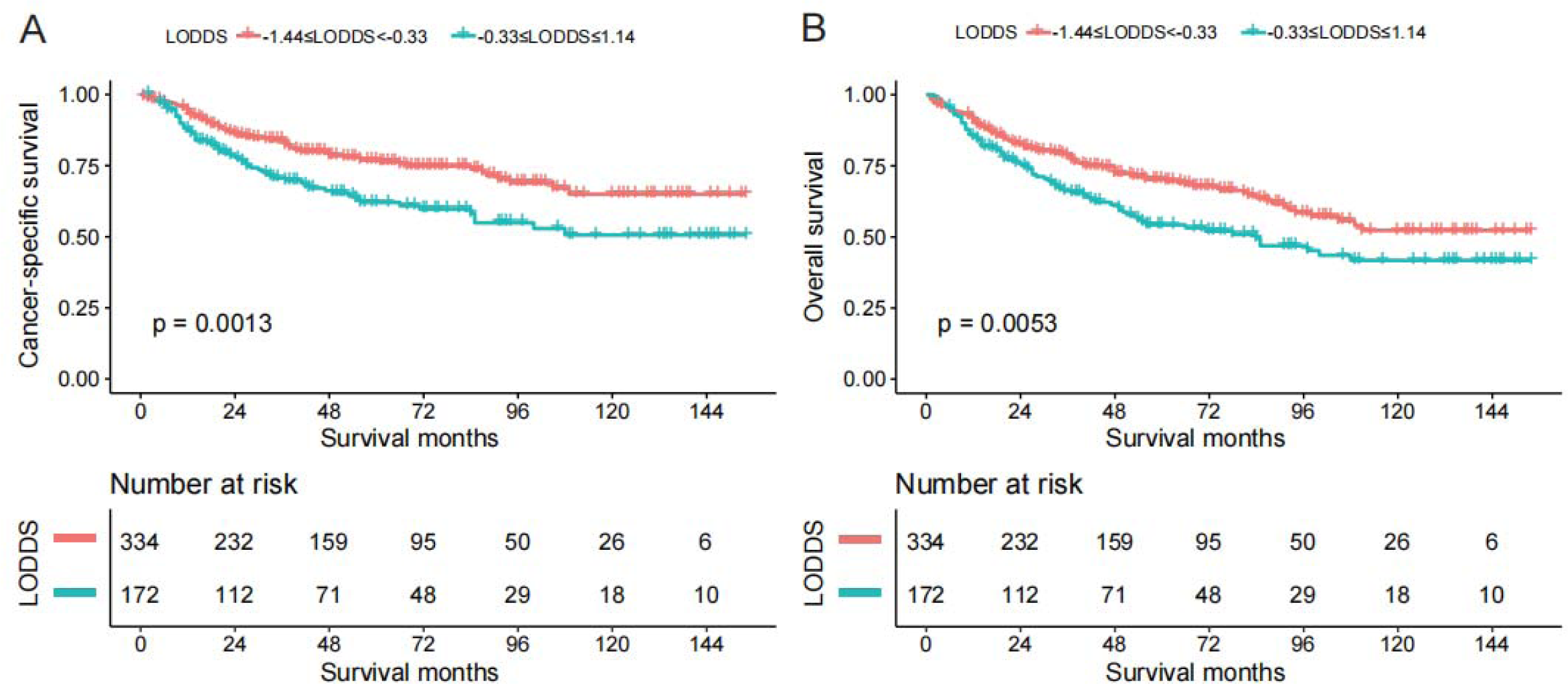
Kaplan-Meier estimates of CSS (A) and OS (B) for entire dataset according to LODDS. CSS: lung cancer-specific survival; OS: overall survival; LODDS: log odds of positive lymph nodes.

**Figure S2.**
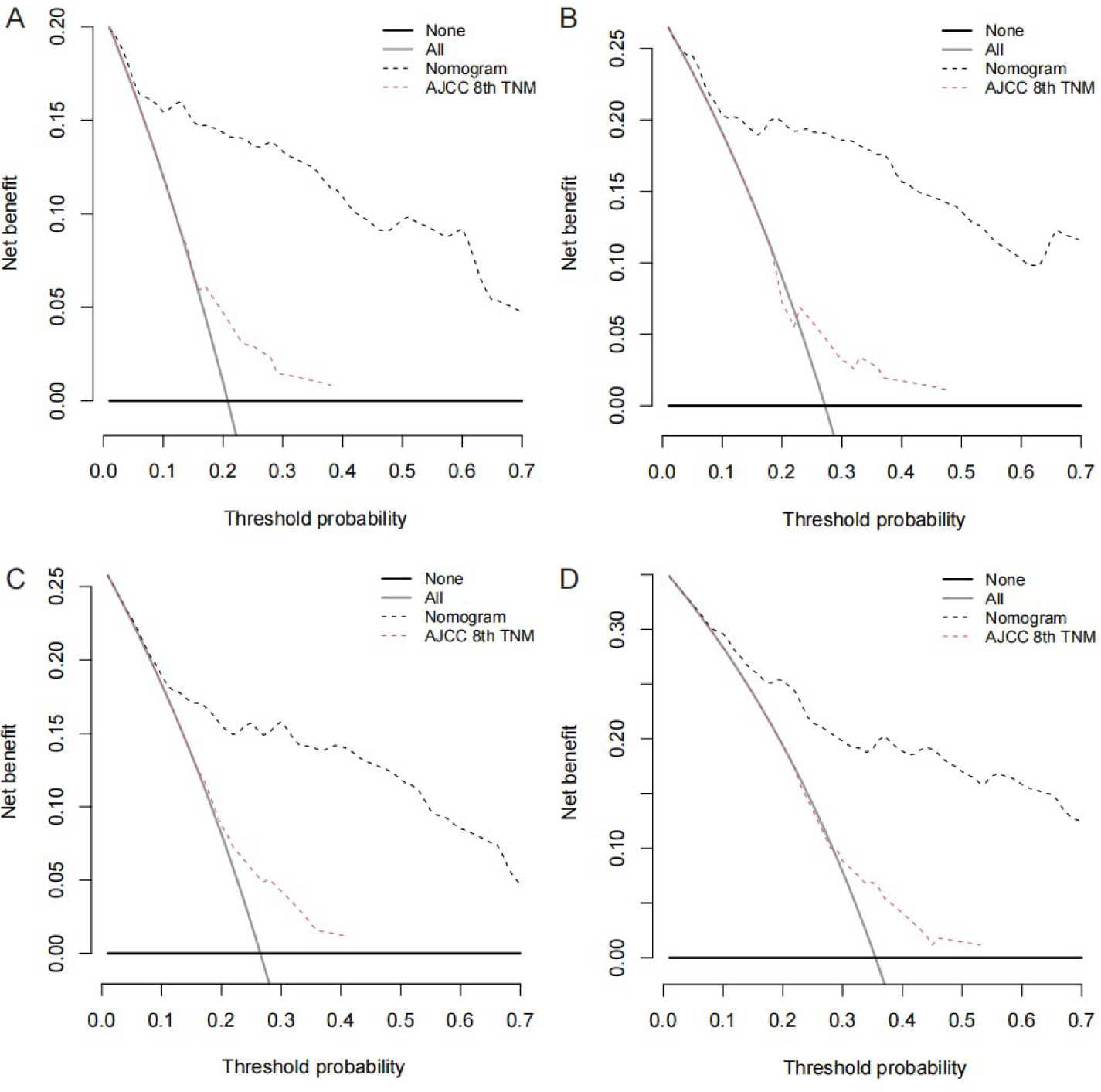
Decision curve analysis of AJCC 8th TNM staging system and nomogram for 3-, and 5-year CSS (A, B), OS (C, D) prediction in the derivation dataset. CSS: lung cancer-specific survival; OS: overall survival; TNM: tumor-node-metastasis.

**Figure S3.**
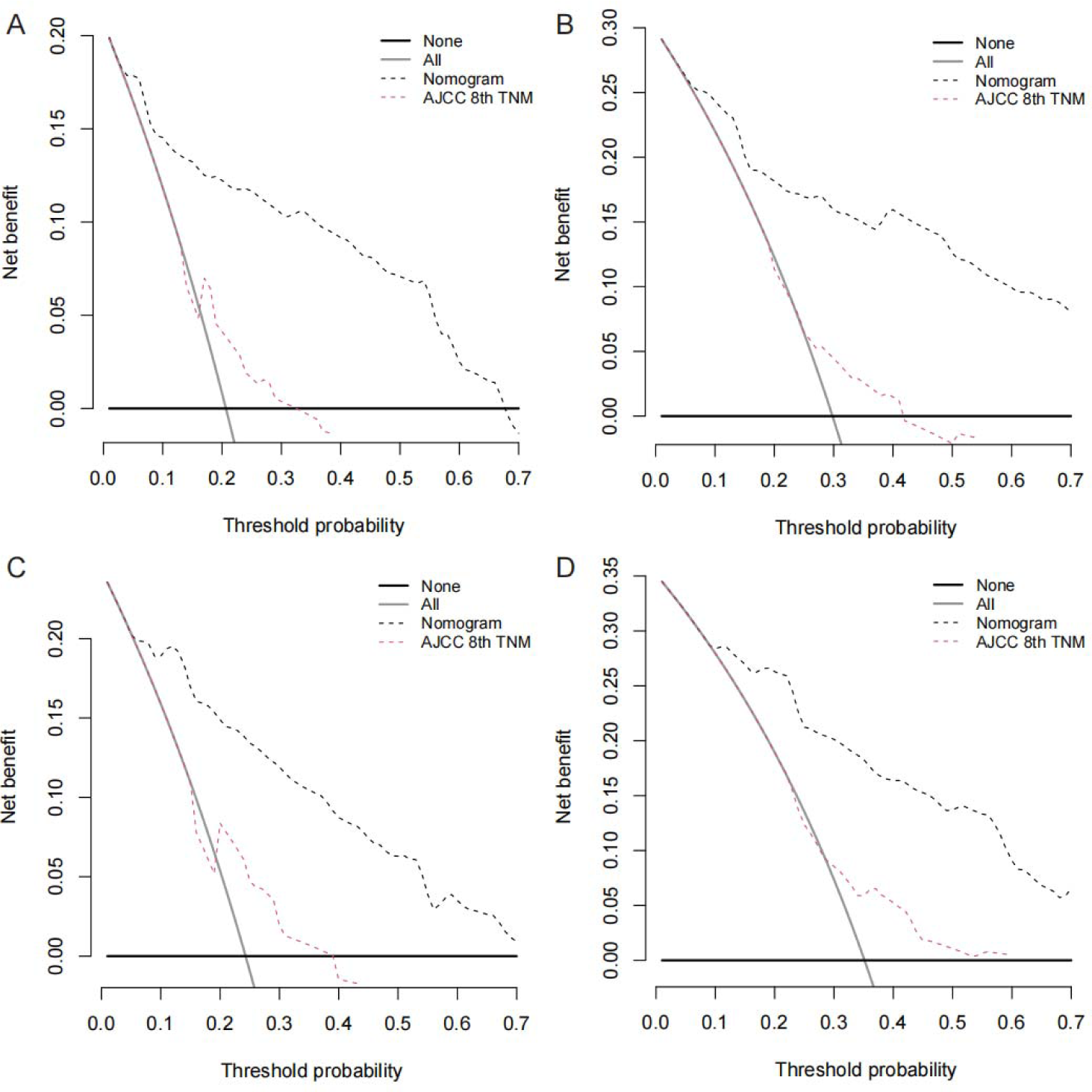
Decision curve analysis of AJCC 8th TNM staging system and nomogram for 3-, and 5-year CSS (A, B), OS (C, D) prediction in the external validation dataset. CSS: lung cancer-specific survival; OS: overall survival; TNM: tumor-node-metastasis.

**Figure S4.**
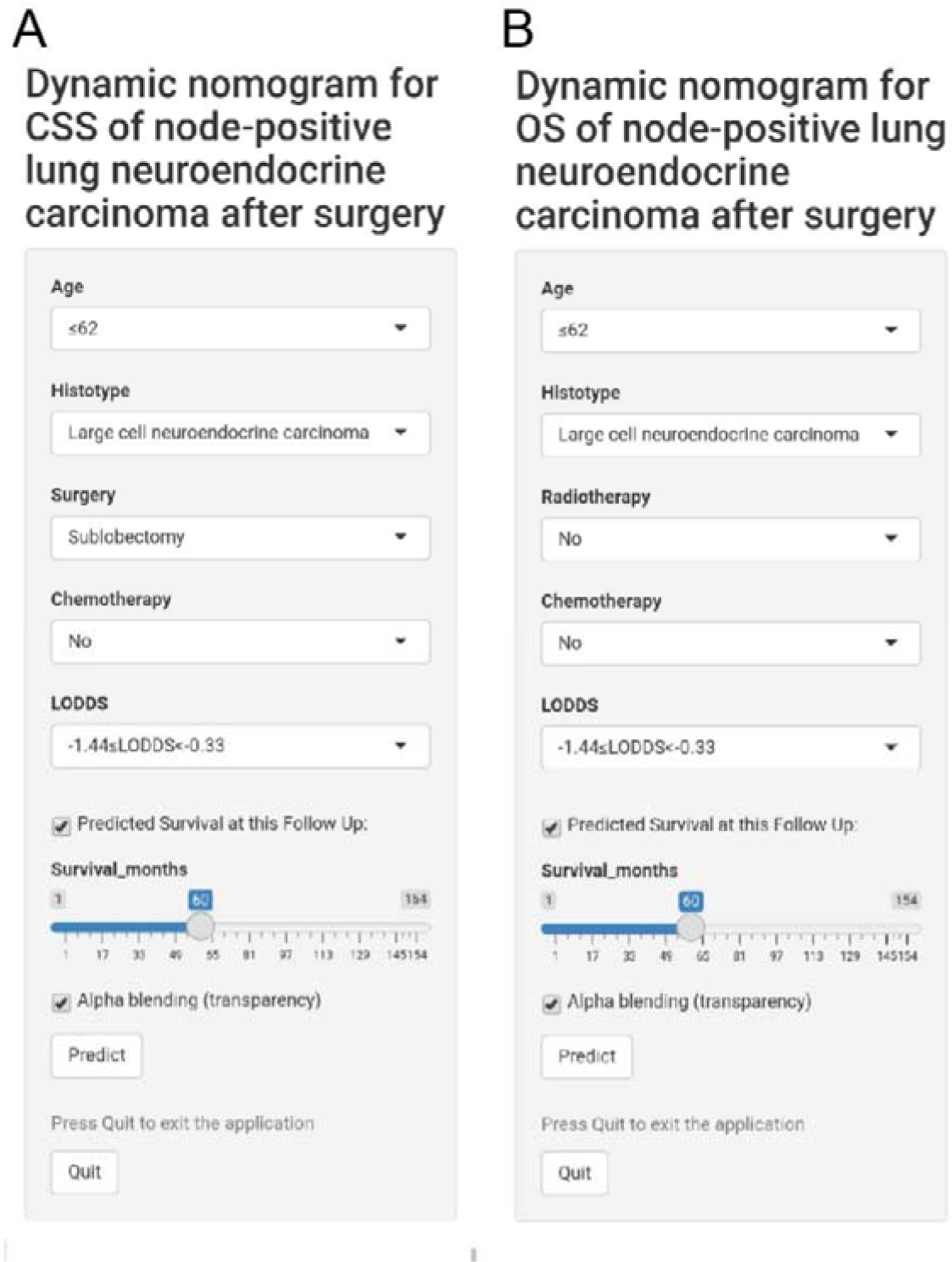
Online nomograms to predict CSS (A) and OS (B) for patients with node-positive lung neuroendocrine carcinoma after surgery. CSS: lung cancer-specific survival; OS: overall survival; LODDS: log odds of positive lymph nodes.

